# BUDS: Benchmark Uncertainty Design Selection for Two-Stage Single-Arm Phase II Trials

**DOI:** 10.64898/2026.05.14.26353210

**Authors:** Rebecca Irlmeier, Zhuoli Jin, Fei Ye

## Abstract

**Background:** Simon two-stage designs for binary endpoints and their time-to-event analogues, including the restricted-Kwak and Jung method, rely on a fixed historical benchmark. In practice, historical benchmarks are often uncertain due to small samples, population heterogeneity, changing eligibility criteria, and evolving standards of care. When the clinically relevant benchmark exceeds the value used to calibrate the design, Type I error rates can be substantially inflated, leading to costly advancement of ineffective treatments. Existing design-selection criteria generally optimize efficiency at a single benchmark without accounting for this uncertainty.

**Methods:** We propose Benchmark Uncertainty Design Selection (BUDS), a framework for selecting two-stage designs for both binary and time-to-event (TTE) endpoints. Rather than relying on a single planning benchmark, BUDS addresses benchmark uncertainty by specifying a plausible range of response rates for binary endpoints or survival probabilities at a prespecified timepoint for TTE endpoints. Among feasible designs that satisfy both Type I error (*α*) and Type II error (*β*) constraints, BUDS-Worst-Regret minimizes the maximum difference in expected sample size from the benchmark-specific optimal design, whereas BUDS-Avg-EN minimizes average expected sample size across the range. By monotonicity, Type I error rate is controlled at the upper bound of the benchmark range.

**Results:** Across representative scenarios, the Type I error rate increases substantially when true benchmarks exceed their planning values using single-benchmark objectives, reaching 0.407 for Simon Optimal design (response rates: 0.10 vs. 0.15; *α* = 0.10, *β* = 0.10) and 0.166 for restricted-Kwak and Jung design (survival probabilities: 0.50 vs. 0.67; *α* = 0.05, *β* = 0.20). In contrast, BUDS-selected designs maintain Type I error rate at or below the nominal level across the benchmark range, while the proposed objectives provide explicit criteria for prioritizing worst-case versus average efficiency.

**Conclusions:** BUDS provides a transparent framework for selecting designs when historical benchmarks are uncertain. It makes the trade-off between robustness and efficiency explicit while controlling Type I error across the plausible benchmark range. The framework applies to both binary and time-to-event endpoints in two-stage single-arm Phase II trial planning and is implemented in the open-source **BUDS** R package and interactive Shiny app.

## Background

Phase II clinical trials serve as a critical gatekeeper in the drug development pipeline (1). Single-arm Phase II trials comparing outcomes against a prespecified historical benchmark remain common in oncology, particularly for rare or biomarker-defined patient subgroups (2).

Classic two-stage designs are calibrated using a single historical benchmark, hereafter referred to as the planning benchmark. For binary endpoints, Simon’s two-stage design has long been widely used due to its operational simplicity and early-termination rules for futility (3). The design selects stagewise boundaries that minimize either the expected (Optimal) or maximum (Minimax) sample size, subject to Type I and Type II error constraints. Subsequent work has considered alternative selection criteria, including graphical search methods for identifying compromised designs, Bayesian decision-theoretic admissible designs based on loss functions, and designs optimized under the alternative hypothesis by minimizing the expected sample size at *p*_1_ (4-6). Analogous two-stage designs have been developed for time-to-event (TTE) endpoints, including methods based on fixed-time survival probabilities and more recent extensions accommodating flexible parametric survival distributions (7-10). Among these, the framework of Kwak and Jung (KJ) and its restricted follow-up time extension by Belin et al. define stagewise test statistics and decision boundaries under a planning null hazard to satisfy prespecified error constraints while minimizing the expected sample size (9, 10).

These existing selection criteria generally evaluate design efficiency at a single fixed benchmark: a response rate *p*_0_ for binary endpoints, or, for TTE endpoints, a survival rate *S*_0_(*x*_0_) at a clinically meaningful time point *x*_0_. In practice, however, these benchmarks are rarely known with certainty (11).

This uncertainty is well illustrated in glioblastoma. For binary endpoints, Phase II temozolomide (TMZ) trials are often designed using Simon’s two-stage framework with a fixed benchmark (12). For example, one current trial assumed *p*_0_ = 0.10, despite observed objective response rates for similar regimens ranging from 5.4% to 13% across studies (13-17). Similarly, a buparlisib trial in recurrent glioblastoma explicitly reported historical 6-month progression-free survival (PFS6) rates of 9% to 16% for ineffective therapies, but used 15% as the null benchmark (17-20).

This benchmark uncertainty reflects sampling variability, differences in patient populations and selection, site-related heterogeneity, endpoint assessment, and temporal changes in clinical practice and supportive care (11,21). Despite these uncertainties, trial designs are typically calibrated using a single benchmark derived from available evidence, implicitly treating it as known (2, 3, 9, 10). In TTE settings, this variability is further compounded by differences in eligibility criteria, prior therapy, follow-up duration, and censoring patterns (17, 20, 22, 23). Consequently, the historical benchmark is often better represented by a range of plausible values than by a single fixed value. The resulting design may therefore be statistically valid at the selected benchmark but insufficiently transparent about why that benchmark, rather than other defensible alternatives, was used.

The fragility of single-arm Phase II designs to benchmark uncertainty is widely recognized. Adaptive and Bayesian designs incorporate uncertainty more formally at the trade-off of operational simplicity or unconditional frequentist guarantees (24-31). With frequentist approaches, underestimating the benchmark inflates the Type I error rate and increases the risk of advancing ineffective treatments; overestimating it reduces statistical power and risks prematurely abandoning potentially effective therapies. A single-arm Phase II design targeting a nominal Type I error rate of 10% can experience an actual false positive rate near 30% when the planning benchmark is underestimated by 5 percentage points from the true response rate of 25% (32). Such benchmark misspecification can lead to incorrect Phase II conclusions, unnecessary later-stage trials, wasted resources, patient exposure to ineffective therapies, and, consequently, its downstream decision reliability.

A key practical gap therefore remains: protocols may acknowledge benchmark uncertainty, but no established framework uses a clinically plausible range to guide prespecified design selection while controlling Type I error across that range. This motivates a two-stage design procedure that formally incorporates benchmark uncertainty during trial planning and selects among feasible designs using an explicit efficiency objective. We address this limitation through Benchmark Uncertainty Design Selection (BUDS), a framework that explicitly incorporates benchmark uncertainty into the design-selection objective. Rather than selecting a two-stage design at a single planning benchmark, BUDS uses a prespecified benchmark range, [*p*_0*L*_,*P*_0*U*_] for binary endpoints or [*S*_0L_,*S*_0*U*_] at a fixed timepoint for TTE endpoints, to make benchmark uncertainty explicit during the trial planning. For each feasible candidate design, a robust objective function is evaluated over the benchmark range under the Type I error constraint. The resulting BUDS-selected designs retain the familiar structure of classic two-stage procedures, yielding stagewise boundaries of the form (*n*_1_,*r*_1_;*n,r*) for binary endpoints and (*n*_1,_*c*_1_,*DA*_1_;*n*_2_,*c*_2_,*DA*_2_) for TTE endpoints. By evaluating candidate designs over a prespecified range of plausible benchmark values, BUDS provides a transparent basis for selecting among feasible two-stage designs. It also quantifies the potential Type I error inflation when the benchmark exceeds the value used to calibrate the design, thereby supporting more transparent trial planning and defensible go/no-go decisions.

## Methods

Two-stage designs are widely used in single-arm Phase II trials to allow for early termination for futility while preserving power to detect promising treatments (33). We consider both binary and TTE endpoints in this two-stage framework.

Let *θ* denote the parameter of interest, representing either the response probability *p* (binary endpoints) or the survival probability *S*(*x*_0_) at a fixed clinically meaningful time point *x*_0_ (TTE endpoints). Let *θ*_0_ and *θ*_1_ represent the null and target alternative parameters, respectively, where *θ*_1_ > *θ*_0_. The formal hypotheses are: H_0_: *θ* ≤ *θ*_0_ vs. H_1_: *θ* > *θ*_0_, with power evaluated at *θ*_1_ and Type I error controlled at *θ*_0_.

### 2.1. Binary Endpoints

We adopt the notation of Simon’s two-stage design and briefly summarize the framework for single-arm Phase II trials with a binary endpoint (3). Let *p* denote the true response probability of the experimental therapy. A design is defined by parameters (*n*_1_, *r*_1_; *n, r*), where *n*_1_ is the sample size of the first stage, *n* is the total planned sample size, and *r*_1_ and *r* are the interim futility and final success boundaries, respectively. Let *X*_1_∼*Binomial(n*_1_,*p*) and *X*_2_∼*Binomial(n*_2_,*p*) denote the number of responses observed in stages 1 and 2. If *X*_1_ ≤ *r*_1_, the trial stops early for futility; otherwise, *n*_2_ = *n* − *n*_1_ additional patients are enrolled in stage 2. The treatment is declared promising (a “Go” decision) if *X* = *X*_1_ + *X*_2_ > *r*.

The probability of declaring success for a given *p*, denoted *α*(*p*), is computed using exact binomial probabilities. Evaluated at the chosen null response probability *p*_0,_ *α*(*p*_0_) corresponds to the Type I error rate; 1 − *β* = *α*(*p*_1_) represents the statistical power under the alternative *p*_1_.

In addition, the probability of early termination (*PET*) and the expected sample size (*EN*) also depend on *p*. Simon designs select (*n*_1_, *r*_1_; *n, r*) to minimize either *n* (Minimax) or *EN*(*p*_0_) (Optimal), subject to Type I and Type II error constraints.

### 2.2. Time-to-Event Endpoints

For TTE outcomes, we build on the two-stage design framework of Kwak and Jung and its restricted follow-up time extension (restricted-Kwak and Jung, or r-KJ) (9, 10). Let *S*(*x*_0_) denote the survival probability at a fixed clinically meaningful time point *x*_0_, modeled under an exponential distribution. Under this formulation, the benchmark and target survival probabilities *S*_0_(*x*_0_) = exp (−*λ*_0_*x*_0_) and *S*_1_(*x*_0_) = exp (−*λ*_1_*x*_0_) correspond to hazard rates *λ*_0_ and *λ*_1_, respectively, and the hazard ratio is defined as HR = *λ*_0_\ *λ*_1_ (consistent with the r-KJ design). Accrual is assumed to be uniform over the interval [0, *t*_*a*_], where *t*_*a*_ is the accrual duration. Follow-up is restricted to *x*_0_, with administrative censoring imposed at that time horizon. *DA*_1_ and *DA*_2_ denote the interim and final analysis calendar times, respectively.

A two-stage design is defined by parameters (*n*_1_, *c*_1_, *DA*_1_; *n*_2_, *c*_2_, *DA*_2_), where *n*_1_ and *n*_2_ are the first-stage and maximum sample sizes and *c*_1_ and *c*_2_ are the corresponding futility boundaries, respectively. Let *Z*_1_ and *Z*_2_ denote the standardized test statistics at the interim and final analyses, arising from stagewise one-sample log-rank tests. Under the assumed exponential hazard model, (*Z*_1_, *Z*_2_) are approximately jointly bivariate normal, with mean and covariance determined by the design parameters and accrual assumptions. If *Z*_1_ > *c*_1_, the trial stops early for futility. Otherwise, *n*_2_ − *n*_1_ additional patients are enrolled and the treatment is declared promising if *Z*_2_ ≤ *c*_2_.

The probability of declaring success *α* (*S*_0_ (*x*_0_)) is obtained from the joint distribution of (*Z*_1_, *Z*_2_). Under the benchmark survival probability, *α* (*S*_0_ (*x*_0_)) defines the Type I error rate, while evaluation at *S*_1_(*x*_0_) yields the corresponding power. Additional operating characteristics, including *PET* and *EN*, also vary with the underlying survival probability. Unlike in the binary setting, the boundaries (*c*_1_,*c*_2_) are continuous and depend explicitly on the survival probability *S*_0_(*x*_0_) through its corresponding hazard rate *λ*_0_. As a result, both the distribution of the test statistics and the resulting operating characteristics vary with the planning benchmark parameter and chosen follow-up horizon *x*_0_.

### 2.3. Benchmark Uncertainty Design Selection (BUDS) Framework

When the control benchmark is uncertain, calibrating a design at a single planning benchmark may not satisfy the Type I error constraint when evaluated at other plausible benchmark values. If the true control rate is higher than anticipated, false positive declarations become more likely, potentially exposing participants to ineffective interventions and misallocating research resources. These considerations motivate incorporating benchmark uncertainty directly into the design-selection objectives. Here, the planning benchmark *θ*_0_ refers to the historical response rate or survival probability used to determine a design’s sample sizes and stopping boundaries. BUDS addresses uncertainty in this value by specifying a plausible range, [*θ*_0*L*_, *θ*_0*U*_]. We specify a clinically plausible benchmark range [*p*_0*L*_, *p*_0*U*_] for binary endpoints, where *p*_0*U*_ < *p*_1_, or [*S*_0*L*_ (*x*_0_), *S*_0*U*_ (*x*_0_)] for TTE endpoints, where *S*_0*U*_ (*x*_0_) < *S*_1_(*x*_0_), and stagewise design boundaries to control the Type I error rate over the entire range. The specification of this benchmark range should be guided by clinical judgement, historical evidence, and the degree of uncertainty surrounding the control benchmark. No universal specification of the benchmark range is assumed.

A BUDS-selected design is defined by parameter set (*n*_1_, *r*_1;_ *n, r*) for binary endpoints and (*n*_1_, *c*_1_, *DA*_1_; *n*_2_, *c*_2_, *DA*_2_) for TTE endpoints. The design parameter set is chosen to satisfy the following constraints:

1. **Type I error control** across the full benchmark range: Pr_*θ*_ (*Go*) ≤ *α*, ∀ *θ* ∈ [*θ*_0*L*_, *θ*_0*U*_]
2. **Power** control at the alternative to detect the desirable treatment effect 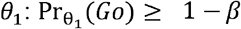
3. **Maximum sample size cap (optional)** to ensure clinical feasibility: *n* ≤ *n*_*max*_

This framework ensures error control over the entire plausible benchmark range, rather than relying on calibration at a single fixed planning value *θ*.

Exact evaluation of the Type I error rate over this range is computationally tractable due to monotonicity. For binary endpoints (Supplementary Material 1, Lemma 1, Corollary 1), both Pr_*p*_(*Go*) and *EN*(*p*) are non-decreasing functions of the true response probability *p*; consequently, the worst-case Type I error and worst-case *EN* within the benchmark range occur at *p*_0*U*_. Type I error control over [*p*_0*L*_, *p*_0*U*_] is therefore equivalent to enforcing the error constraint at *p*_0*U*_ alone. Analogously, for TTE endpoints, monotonicity properties hold with respect to the survival probability *S*(*x*_0_), and thus Type I error rate is evaluated at the boundary *S*_0*U*_ (*x*_0_). Formal proofs of these properties are provided in the Supplementary Material 2.

### 2.4. BUDS Objectives

A key feature of BUDS is that design selection optimizes a robust objective defined over the entire plausible benchmark range, rather than at a single planning benchmark, as is commonly done in practice.

Our primary objective, **BUDS-Worst-Regret**, minimizes the maximum regret across the plausible benchmark range. For any candidate design *d* and any value *θ* ∈ [*θ*_0*L*_, *θ*_0*U*_], regret is defined as:

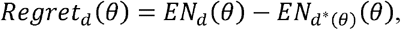

where

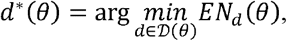

and *D*(*θ*) is the set of designs satisfying the Type I error and power constraints when the benchmark is *θ*. Thus, *d*_*_(0) is the locally optimal design that would be selected if *θ* were known. BUDS-Worst-Regret selects

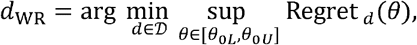

where

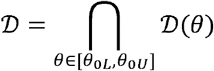

is the set of designs satisfying the operating-characteristic constraints throughout the benchmark range. This objective provides a principled hedge against benchmark uncertainty by selecting the feasible design whose expected sample size deviates least, in the worst case, from the benchmark-specific optimum. The existence of *Regret*_*d*_ (*θ*) and d_*WR*_ for binary endpoints is established in Supplementary Material 1 (Proposition 2 and 3); the same argument extends to TTE endpoints.

Alternatively, the **BUDS-Avg-EN** objective minimizes the average *EN* across the benchmark range, computed over a fine grid of *θ* values spanning [*θ*_0*L*_, *θ*_0*U*_]. This objective favors designs that are efficient on average across the prespecified benchmark range.

### 2.5. BUDS Algorithms

Designs selected under the proposed BUDS objectives are obtained via computationally efficient search algorithms that differ fundamentally between binary and TTE endpoints.

For binary endpoints, the design space *D* can be exhaustively enumerated over all candidate two-stage configurations. The search space contains 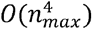 candidate parameter combinations; vectorization and early pruning are used to reduce computational burden.

For TTE endpoints, candidate designs are constructed using a search procedure based on the r-KJ framework, with feasibility constraints enforced throughout. For each benchmark survival probability in the prespecified benchmark range, a localized “zoom” search is conducted over (*n*_1_, *c*_1_,*n*_2_), initialized from the corresponding single-stage design. The final-stage boundary *c*_2_ is determined to satisfy both Type I and Type II error constraints. In the current implementation, the grid is evaluated at increments of 0.01. Candidate designs retained across benchmark values are subsequently pooled and evaluated.

The general algorithm proceeds as follows:

#### Step 1. Generate Candidate Designs and Enforce the Power Constraint

Specify a maximum sample size cap, *n*_max_, and define fine grids *G*_*p*_ and *G*_*s*_ spanning [*p*_0*L*,_ *p*_0*U*_] and [*S*_0*L*_(*x*_0_), *S*_0*U*_ (*x*_0_)] respectively.

- **Binary:** Enumerate all integer parameter sets (*n*_1_, *r*_1_, *n, r*) satisfying 1 ≤ *n*_1_ < *n* ≤ *n*_max_, 0 ≤ *r*_1_ ≤ *n*_1_ and *r*_1_ < *r* < *n*. For each candidate design, compute Pr _*p*1_(G0) using exact binomial probabilities. Retain designs satisfying

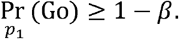
- **TTE:** For each calibration benchmark 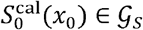. where 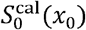 denotes the null survival probability used to calibrate the design, generate candidate designs through a “zoom” search over (*n*_1_, *c*_1_,*n*_2_ ), initialized from the corresponding single-stage KJ design and subject to *n*_2_ ≤ *n*_max_. At each iteration, determine *c*_2_ to satisfy the local Type I error constraint

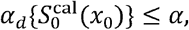

and retain designs achieving the required power

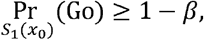

with *S*_1_(*x*_0_) held fixed.

#### Step 2. Enforce Type I Error Control Across the Benchmark Range

For each candidate design retained in Step 1:

- **Binary:** Evaluate the worst-case Type I error at *p*_0*U*_. Retain designs satisfying

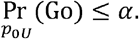
- **TTE:** With the decision boundaries fixed at those determined under 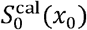, use closed-form expressions to evaluate Type I error over *G*_*s*_. Retain designs satisfying

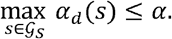

**Step 3. Identify the Feasible Design Set**

- **Binary:** Let *D* denote the set of designs retained after Step 2.
- **TTE:** Pool the designs retained across all calibration benchmarks 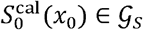. and let *D* denote the resulting feasible design set.

#### Step 4. Evaluate the Optimization Objective

For each *d* ∈ *D*, evaluate the selected robust objective, BUDS-Worst-Regret or BUDS-Avg-EN, over the corresponding benchmark grid.

- **Binary:** Evaluate each design over *p* ∈*G*_*p*_ and select the design minimizing the chosen objective.
- **TTE:** Evaluate each design over *s* ∈ *G*_*s*_ and select the design minimizing the chosen objective. The reported 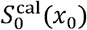 identifies the benchmark used to construct the selected design.

#### Step 5. Apply Tie-Breaking Rules

If multiple designs minimize the objective in Step 4, select the design with the smaller maximum sample size - *n* for binary endpoints or *n*_2_ for TTE endpoints. If a tie persists, select the design with the smaller average *EN* across the corresponding benchmark grid.

These algorithms have been implemented in the open-source R package, **BUDS**, with C++ implementation via Rcpp (34) for computational efficiency. The package provides functions for selecting designs that optimize the proposed robust objectives and computing exact operating characteristics. An interactive Shiny application has been developed to allow users to specify design parameters, incorporate benchmark uncertainty, and visualize operating characteristics and design trade-offs in real time. Both the R package and Shiny tool are available on GitHub: https://github.com/Ye-Lab-UMiami/BUDS.

## Results

To evaluate the impact of historical benchmark uncertainty on two-stage designs, we compare designs calibrated at a single planning benchmark with BUDS-selected designs for binary and TTE endpoints. Simon and r-KJ designs are calibrated at both *θ*_0_ and the upper bound of the benchmark range, *θ*_0*U*_: the former illustrates the consequences of benchmark misspecification, whereas the latter holds the feasible design set fixed and directly compares the selection criteria. Comparisons focus on three key aspects: (i) robustness to benchmark misspecification, assessed by the Type I error rate across [*θ*_0*L*_, *θ*_0*U*_], (ii) design efficiency, assessed by expected and maximum sample sizes, and (iii) design-performance trade-offs across benchmark ranges and selection objectives.

The primary binary endpoint results are evaluated using one-sided nominal error rates of (*α,β*) = (0.10, 0.10). Three representative settings are considered: (*p*_0_, *p*_1_) = (0.10, 0.25), (0.20, 0.40), and (0.30, 0.50), reflecting a range of response rates commonly encountered in oncology trials. Additional binary endpoint results using (*α,β*) = (0.05, 0.20) are provided in Supplementary Material 3 (Table S1).

For TTE endpoints, the primary results are evaluated using (*α, β*) = (0.05, 0.20). Scenarios are defined through survival probability at a clinically meaningful time point *x*_0_ = 1. We consider (*S*_0_(*x*_0_), *S*_1_(*x*_0_)) = (0.15, 0.34), (0.35, 0.59), and (0.50, 0.67). A constant accrual rate of 30 patients per unit time is assumed. Additional TTE results using (*α,β*) = (0.10, 0.10) are provided in Supplementary Material 3 (Table S2).

For both endpoint types, increasing degrees of benchmark uncertainty are represented through clinically plausible benchmark ranges, [*θ*_0*L*_, *θ* _0*U*_]:

- **Single benchmark value:** [*θ* _0_, *θ*_0_], corresponding to the classic binary (Simon) and TTE (r-KJ) designs
- **Moderate benchmark uncertainty:** [*θ*_0_ − 0.02, *θ*_0_ + 0.02]
- **Substantial benchmark uncertainty:** [*θ*_0_ − 0.05, *θ*_0_ + 0.05]

where *θ*_0_ denotes the benchmark value (response probability for binary endpoints and survival probability at *x*_0_ for TTE endpoints). The qualitative labels of these configurations are used solely for reporting convenience. Unless otherwise noted, results reporting focuses on designs selected by the BUDS-Worst-Regret objective. Findings for BUDS-Avg-EN are summarized separately in Section 3.4.

### 3.1. Robustness to Benchmark Misspecification

We first examine Type I error behavior when designs are evaluated at benchmark values that differ from the planning benchmark. Table 1 summarizes the design boundaries and key operating characteristics of designs for binary endpoints selected by the Simon Optimal, Simon Minimax, and BUDS-Worst-Regret objectives under increasing degrees of uncertainty. Table 2 presents the corresponding results for TTE endpoints using the r-KJ and BUDS-Worst-Regret objectives. Figure 1 illustrates this behavior by displaying the exact Type I error rate as a function of the benchmark value used for evaluation across the plausible range.

**Table 1.**
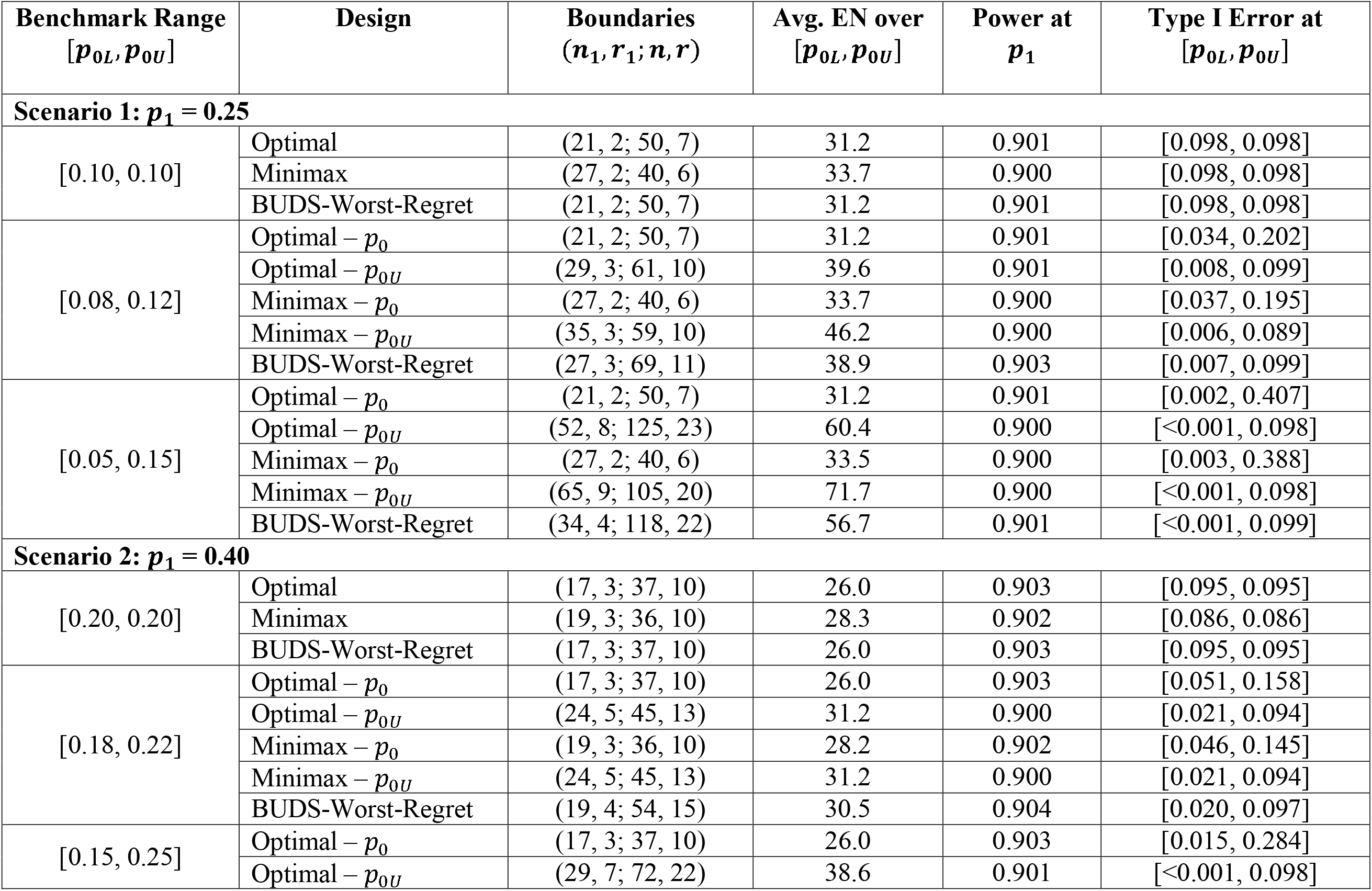
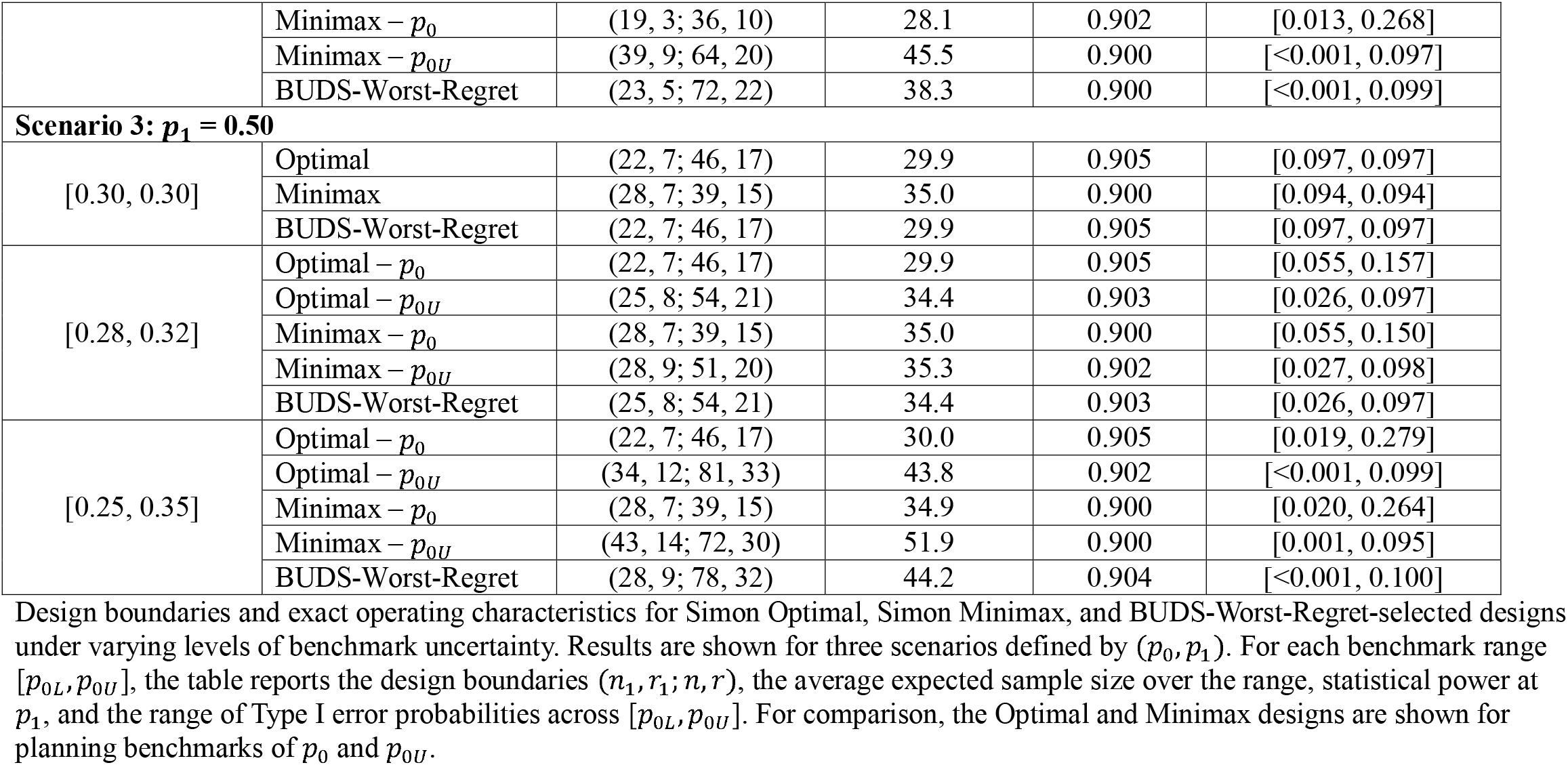
Design Boundaries and Exact Operating Characteristics Under Benchmark Uncertainty for Binary Endpoints (α = 0.10, β = 0.10, n_max_ = 150)

**Table 2.** Design Boundaries and Exact Operating Characteristics Under Benchmark Uncertainty for Time-to-Event Endpoints (α = 0.05, β = 0.20, n_max_ = 150)

| Benchmark Range<br>$[S_{0L}(x_0), S_{0U}(x_0)]$ | Design | Calibration<br>Benchmark<br>$S_0^{\text{cal}}(x_0)$ | Boundaries<br>$(n_1, c_1, DA_1; n_2, c_2, DA_2)$ | Avg. EN over<br>$[S_{0L}(x_0), S_{0U}(x_0)]$ | Power at<br>$S_1(x_0)$ | Type I error at<br>$[S_{0L}(x_0), S_{0U}(x_0)]$ |
| --- | --- | --- | --- | --- | --- | --- |
| **Scenario 1: $S_1(x_0) = 0.34$** |  |  |  |  |  |  |
| [0.15, 0.15] | r-KJ | 0.15 | (18, 0.37, 0.58; 27, -1.62, 1.90) | 23.8 | 0.801 | [0.050, 0.050] |
|  | BUDS-Worst-Regret | 0.15 | (18, 0.37, 0.58; 27, -1.62, 1.90) | 23.8 | 0.801 | [0.050, 0.050] |
| [0.13, 0.17] | r-KJ $- S_0(x_0)$ | 0.15 | (18, 0.37, 0.58; 27, -1.62, 1.90) | 23.8 | 0.801 | [0.027, 0.086] |
| | r-KJ $- S_{0U}(x_0)$ | 0.17 | (24, 0.17, 0.78; 36, -1.62, 2.20) | 29.9 | 0.801 | [0.012, 0.050] |
|  | BUDS-Worst-Regret | 0.17 | (24, 0.21, 0.80; 36, -1.63, 2.20) | 30.1 | 0.806 | [0.012, 0.050] |
| [0.10, 0.20] | r-KJ $- S_0(x_0)$ | 0.15 | (18, 0.37, 0.58; 27, -1.62, 1.90) | 23.8 | 0.801 | [0.009, 0.169] |
| | r-KJ $- S_{0U}(x_0)$ | 0.20 | (36, 0.05, 1.18; 57, -1.63, 2.90) | 42.6 | 0.801 | [<0.001, 0.050] |
|  | BUDS-Worst-Regret | 0.20 | (29, 0.21, 0.96; 62, -1.61, 3.07) | 42.7 | 0.800 | [<0.001, 0.050] |
| **Scenario 2: $S_1(x_0) = 0.59$** |  |  |  |  |  |  |
| [0.35, 0.35] | r-KJ | 0.35 | (19, 0.42, 0.61; 28, -1.62, 1.93) | 25.0 | 0.802 | [0.050, 0.050] |
|  | BUDS-Worst-Regret | 0.35 | (19, 0.42, 0.61; 28, -1.62, 1.93) | 25.0 | 0.802 | [0.050, 0.050] |
| [0.33, 0.37] | r-KJ $- S_0(x_0)$ | 0.35 | (19, 0.42, 0.61; 28, -1.62, 1.93) | 25.0 | 0.802 | [0.034, 0.073] |
| | r-KJ $- S_{0U}(x_0)$ | 0.37 | (22, 0.26, 0.72; 34, -1.62, 2.13) | 28.7 | 0.801 | [0.020, 0.050] |
|  | BUDS-Worst-Regret | 0.37 | (23, 0.21, 0.76;<br>34, -1.62, 2.13) | 28.9 | 0.803 | [0.020, 0.050] |
| [0.30, 0.40] | r-KJ – $S_0(x_0)$ | 0.35 | (19, 0.42, 0.61;<br>28, -1.62, 1.93) | 24.9 | 0.802 | [0.018, 0.122] |
| | r-KJ – $S_{0U}(x_0)$ | 0.40 | (30, 0.16, 0.97;<br>45, -1.62, 2.50) | 36.4 | 0.801 | [0.003, 0.050] |
|  | BUDS-Worst-Regret | 0.40 | (30, 0.21, 1.00;<br>45, -1.63, 2.50) | 36.6 | 0.806 | [0.003, 0.050] |
| **Scenario 3: $S_1(x_0) = 0.67$** |  |  |  |  |  |  |
| [0.50, 0.50] | r-KJ | 0.50 | (36, 0.01, 1.19;<br>57, -1.62, 2.90) | 46.5 | 0.800 | [0.050, 0.050] |
|  | BUDS-Worst-Regret | 0.50 | (36, 0.01, 1.19;<br>57, -1.62, 2.90) | 46.5 | 0.800 | [0.050, 0.050] |
| [0.48, 0.52] | r-KJ – $S_0(x_0)$ | 0.50 | (36, 0.01, 1.19;<br>57, -1.62, 2.90) | 46.6 | 0.800 | [0.029, 0.084] |
| | r-KJ – $S_{0U}(x_0)$ | 0.52 | (44, -0.03, 1.46;<br>72, -1.62, 3.40) | 55.8 | 0.800 | [0.013, 0.050] |
|  | BUDS-Worst-Regret | 0.52 | (48, -0.21, 1.60;<br>72, -1.62, 3.40) | 56.3 | 0.800 | [0.013, 0.050] |
| [0.45, 0.55] | r-KJ – $S_0(x_0)$ | 0.50 | (36, 0.01, 1.19;<br>57, -1.62, 2.90) | 46.6 | 0.800 | [0.012, 0.166] |
| | r-KJ – $S_{0U}(x_0)$ | 0.55 | (65, -0.07, 2.15;<br>109, -1.62, 4.63) | 77.4 | 0.800 | [<0.001, 0.050] |
|  | BUDS-Worst-Regret | 0.55 | (55, 0.07, 1.82;<br>117, -1.61, 4.90) | 77.6 | 0.800 | [<0.001, 0.050] |

Design boundaries and exact operating characteristics for restricted-KJ and BUDS-Worst-Regret-selected designs under varying levels of benchmark uncertainty. Results are shown for three scenarios defined by $(S_0(x_0), S_1(x_0))$ . For each benchmark range $[S_{0L}(x_0), S_{0U}(x_0)]$ , the table reports the design boundaries $(c_1, n_1, DA_1; c_2, n_2, DA_2)$ , the average expected sample size over the range, statistical power at $S_1(x_0)$ , and the range of Type I error probabilities across $[S_{0L}(x_0), S_{0U}(x_0)]$ . The column *Calibration Benchmark* $S_0^{cal}(x_0)$ denotes the evaluation benchmark survival probability at which the design selected by the objective is constructed. For comparison, the r-KJ design is shown for planning benchmarks of $S_0(x_0)$ and $S_{0U}(x_0)$ , with the target survival probability $S_1(x_0)$ ) held fixed in both cases (resulting in different derived hazard ratios). Design inputs: $x_0 = 1$ , accrual rate = 30.

**Figure 1:**
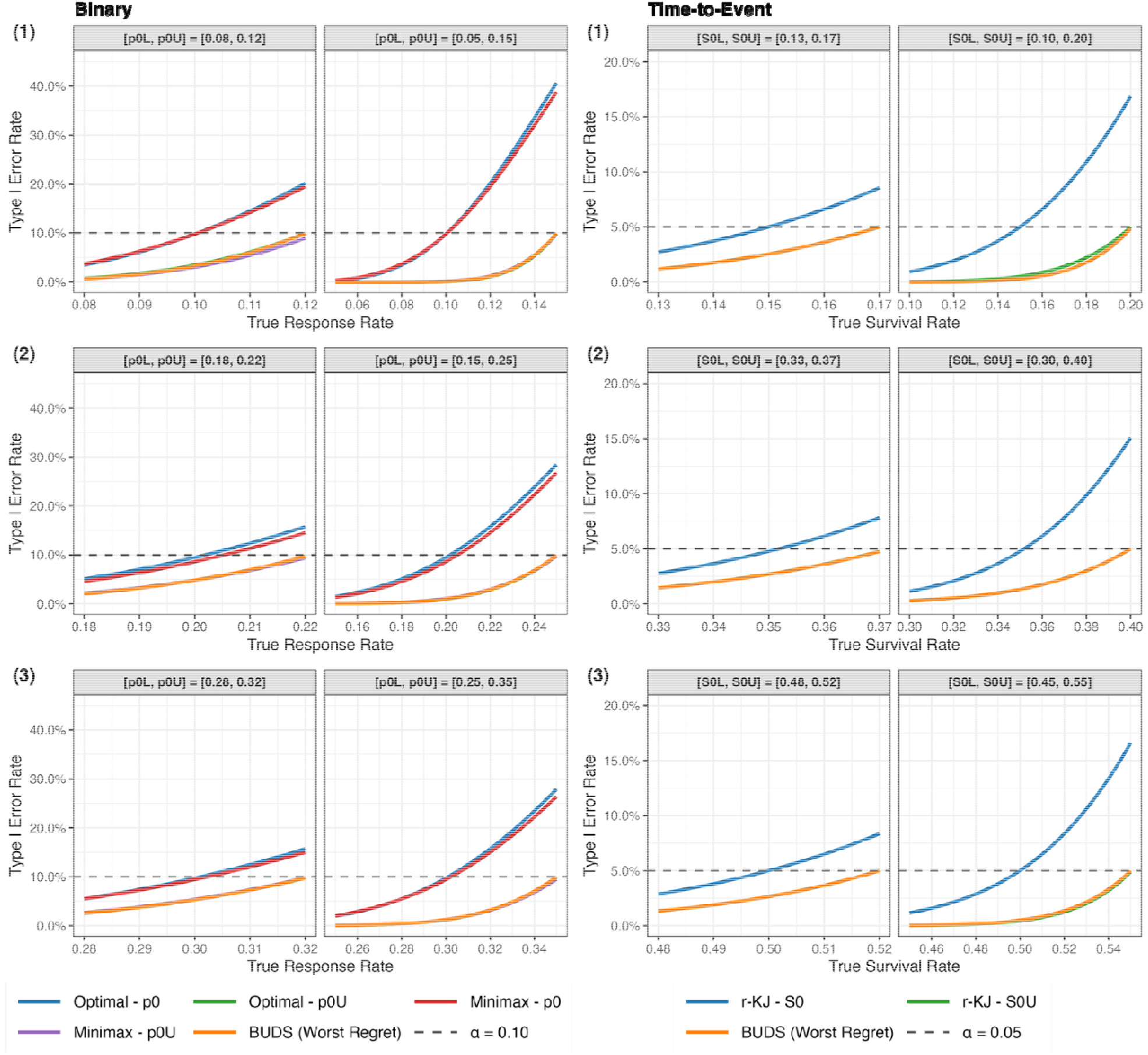
Type I Error Rate as a Function of the True Benchmark by Design. **Legend:** Type I error curves for designs selected by single-benchmark and BUDS objectives across the benchmark range. Panel (A) shows binary endpoint scenarios with = (0.10, 0.25), (0.20,0.40), and (0.30, 0.50), planned with = 0.10, = 0.10, and = 150. Panel (B) shows time-to-event scenarios defined by ) = (0.15,0.34), (0.35,0.59), and (0.50, 0.67) at = 1,planned with =0.05, = 0.20, a uniform accrual rate of 30, and = 150. The dashed horizontal lines denote the nominal Type I error level. “Within each scenario, results are shown for two benchmark ranges, representing moderate and substantial uncertainty. For comparison, the single-benchmark designs are shown for planning benchmarks of both and . The BUDS-Worst-Regret-selected designs maintain Type I error rate at or below throughout the benchmark range, whereas classic designs calibrated at exhibit increasing Type I error rate as the evaluation benchmark approaches the upper boundary of the range. Because Type I error is monotone in the benchmark, designs calibrated at also control Type I error across the benchmark range; consequently, some BUDS and -calibrated designs coincide.

In the binary setting under substantial uncertainty, the Simon Optimal design calibrated at *p*_0_ = 0.10 (Scenario 1) has a Type I error rate of 0.407 when evaluated at the upper boundary of the benchmark range - more than four times the nominal 10% level. Similarly, in the TTE setting, when the true baseline survival exceeds the planning benchmark, the r-KJ design can yield an inflated type I error rate. This is illustrated in Scenario 3 under substantial uncertainty around *S*_0_(*x*_0_) = 0.50, where the Type I error rate increases to approximately 0.17, more than three times the nominal *α* = 0.05. For both endpoints, BUDS incorporates the benchmark range directly into the design-selection objective. As a result, the selected designs maintain the Type I error rate at or below *α* for every value within the selected benchmark range, including the worst-case configuration.

### 3.2. Design Efficiency

We next examine the efficiency implications of enforcing Type I error control across a range of plausible benchmark values. While BUDS guarantees validity under benchmark uncertainty, this robustness requires additional information, reflected in the increased sample size of the resulting design. Figure 2 compares the first-stage, expected, and maximum sample sizes of single-benchmark and BUDS-selected designs.

**Figure 2:**
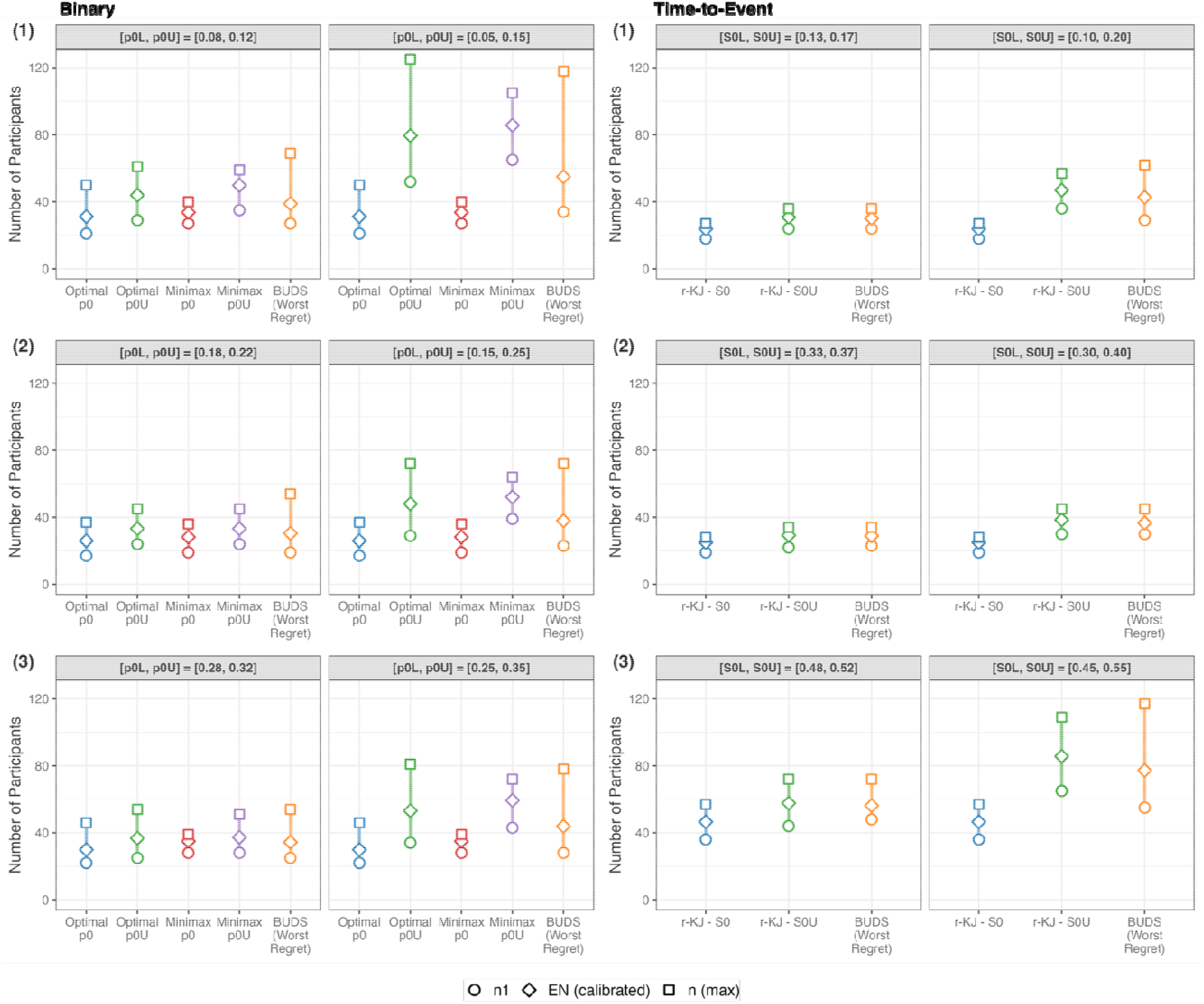
First-Stage, Expected, and Maximum Sample Sizes by Design. **Legend:** Panel (A) shows binary endpoint scenarios with = (0.10, 0.25), (0.20, 0.40), and (0.30, 0.50), planned with = 0.10, = 0.10, and = 150. Panel (B) shows time-to-event scenarios defined by ) = (0.15, 0.34), (0.35, 0.59), and (0.50, 0.67) at = 1, planned with = 0.05, = 0.20, a uniform accrual rate of 30, and = 150. Within each scenario, results are shown for two benchmark ranges, representing moderate and substantial uncertainty. For comparison, the single-benchmark designs are shown for planning benchmarks of both and . Symbols denote the first-stage sample size, the expected sample size evaluated at the benchmark value used to calibrate each design ( or ), and the maximum sample size. As the benchmark range widens, the BUDS-Worst-Regret-selected design requires a larger maximum sample size, reflecting the additional information needed to preserve power while enforcing Type I error control at the least favorable benchmark configuration.

Under single-benchmark specification, the BUDS-Worst-Regret objective selects the design that coincides with the corresponding classic design. These are ] for Simon Optimal and Minimax designs (Table 1) and for r-KJ design (Table 2). As uncertainty in the benchmark is introduced, however, the maximum sample size increases accordingly. For the binary endpoint, widening the benchmark range yields nested feasible sets (Supplementary Material 1, Proposition 1). Consequently, the minimum feasible maximum sample size is nondecreasing with range width (Supplementary Material 1, Corollary 3). For the TTE endpoint, the same conclusion follows by analogous arguments. Widening the benchmark range restricts the feasible set, while the worst-case *EN* occurs at the upper bound (Supplementary Material 1, Corollary 2 for binary endpoints; analogous derivation for TTE endpoints).

This pattern holds for both endpoint types when comparing the BUDS-Worst-Regret objective with the design selected by the corresponding single-benchmark objective at *θ*_0_. For example, in the binary Scenario 2 (*p*_0_ = 0.20 *p*_1_ = 0.40), the maximum sample size increases from 37 under the single-benchmark specification to 54 and 72 under moderate and substantial uncertainty, respectively. Likewise, in TTE Scenario 1 (*S*_0_ (*x*_0_) = 0.15, *S*_1_(*x*_0_) = 0.34), the maximum sample size increases from 27 to 36 to 62. These increases reflect the additional enrollment required to distinguish the alternative from the least favorable benchmark configuration while maintaining the desired Type I error control.

### 3.3. Design Performance Trade-offs

Figure 3 summarizes the robustness and efficiency trade-offs by jointly displaying expected and maximum sample sizes while identifying designs that preserve Type I error control across the benchmark range. For both binary and TTE endpoints, narrower benchmark ranges generally yield smaller designs, whereas wider benchmark ranges for robustness to benchmark misspecification generally require additional enrollment.

**Figure 3:**
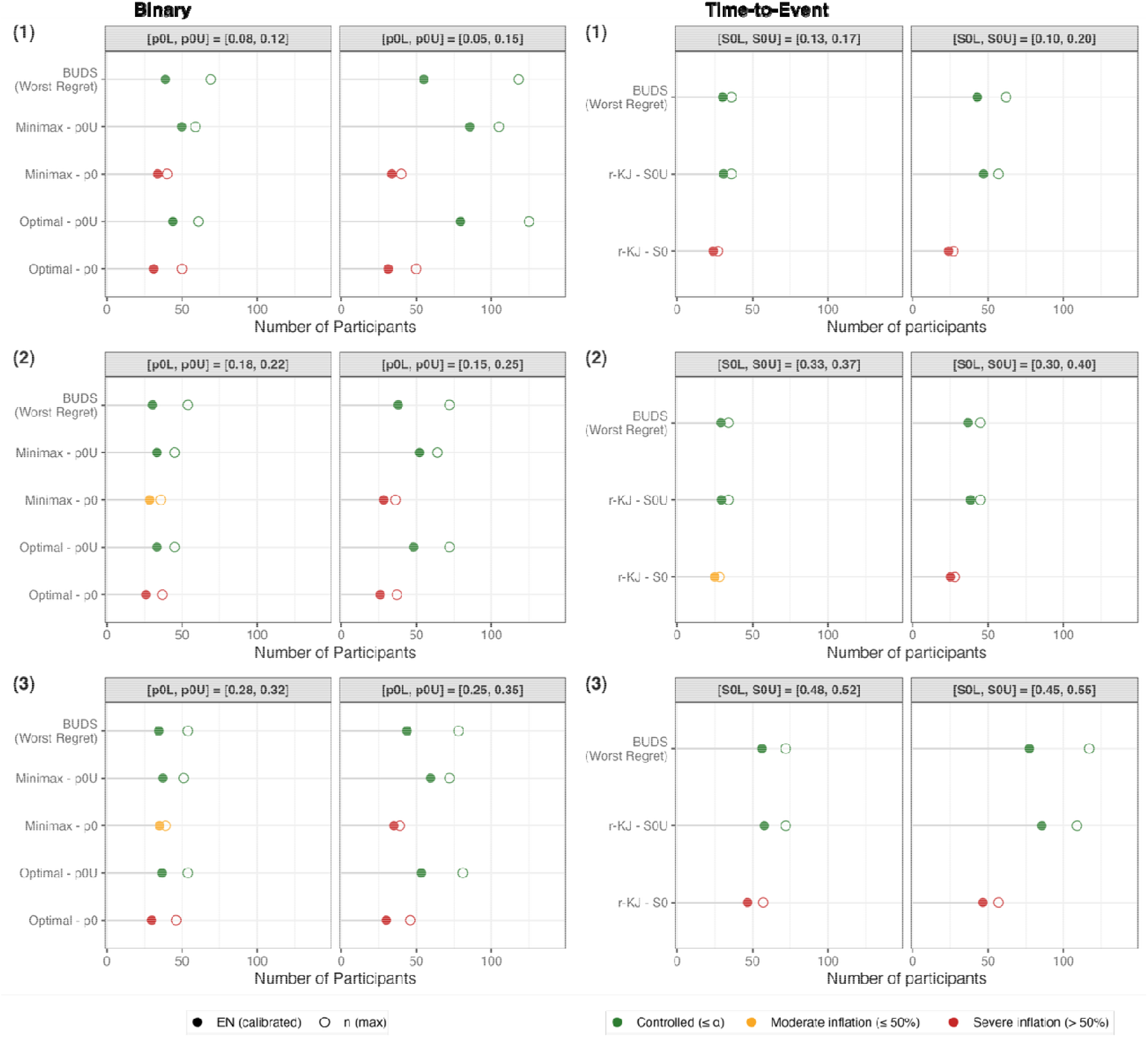
Sample Size and Type I Error Control Under Benchmark Uncertainty. **Legend:** Panel (A) shows binary endpoint scenarios with = (0.10, 0.25), (0.20, 0.40), and (0.30, 0.50), planned with = 0.10, = 0.10, and = 150. Panel (B) shows time-to-event scenarios defined by ) = (0.15, 0.34), (0.35, 0.59), and (0.50, 0.67) at = 1, planned with = 0.05, = 0.20, a uniform accrual rate of 30, and = 150. Within each scenario, results are shown for two benchmark ranges, representing moderate and substantial uncertainty. For comparison, the single-benchmark designs are shown for planning benchmarks of both and . Symbols denote the expected sample size (filled circle) evaluated at the benchmark value used to calibrate each design ( or ), and the maximum sample size (open circle). Colors indicate whether Type I error is controlled ( )or exhibits inflation. Across both endpoint types, the figure illustrates the trade-off between robustness and efficiency: as the benchmark range widens, BUDS-Worst-Regret accepts an increase in sample size to preserve Type I error control.

Tables 1, 2, S1, and S2, and Figures 1-3 also include designs selected by the single-benchmark objectives at *θ*_0*U*_, all of which maintain Type I error control across the benchmark range. For binary endpoints, the BUDS-Worst-Regret objective often selects designs with maximum sample sizes at or between those of the corresponding Simon Minimax and Simon Optimal designs. These designs generally achieve lower average EN across the benchmark range than Simon Minimax and smaller first-stage sample sizes than either Simon design. For TTE endpoints, the BUDS-Worst-Regret and r-KJ designs show similar sample size trade-offs. As benchmark uncertainty increases, BUDS-Worst-Regret often selects designs with a smaller first-stage sample size, accompanied by a modest increase in maximum sample size, while maintaining a comparable average EN across the benchmark range.

### 3.4 BUDS-Avg-EN Designs

Although the results presented in this paper focus primarily on BUDS-Worst-Regret, we also evaluated BUDS-Avg-EN as an alternative design-selection objective. Across considered scenarios, the two objectives frequently selected similar designs, as both optimize *EN* behavior over the same benchmark range, and *EN* varies monotonically with the underlying parameter for two-stage designs (Supplementary Material 1, Lemma 2). This behavior is observed for both binary and TTE endpoints. Differences arise when candidate designs yield comparable average *EN* but differ in their behavior near the upper boundary of the benchmark range, where worst-case behavior occurs. For example, in the binary setting under Scenario (1) with *p*_0_ = 0.10, *p*_1_ = 0.25, [*p*_0*L*,_, *p*_0*U*_] = [0.05, 0.15], and error rates (*α, β*) = (0.10, 0.20), BUDS-Avg-EN selects (*η*_1_, *r*_1;_ *n, r*) = (24, 3; 84, 16), *EN*(*p*_0*U*_) - 53.7, whereas BUDS-Worst-Regret selects (23, 3; 96, 18), *EN*(*p*_0*U*_) = 56.6. In this setting, the BUDS-Avg-EN objective favors slightly greater overall efficiency, while the BUDS-Worst-Regret objective accepts modestly greater average enrollment to limit the largest efficiency loss relative to the benchmark-specific optimum. Type I error curves for BUDS-Avg-EN designs are provided in Supplementary Material 3 (Figure S1).

### 3.5 Real Trial Applications

To illustrate the practical implications of incorporating benchmark uncertainty into the design-selection process, we apply BUDS to the recurrent glioblastoma Phase II trial settings for both binary and TTE endpoints (18, 35).

For binary endpoints, consider the single-arm Phase II trial of TMZ-based therapy designed using Simon’s two-stage framework with *p*_0_ = 0.10, *p*_1_ = 0.25, *α* =0.10, and *β* = 0.10. Under this specification, the Simon Optimal and Minimax designs yield maximum sample sizes of 50 and 40, respectively. However, when incorporating a plausible benchmark range from literature, [*p*_0*L*_, *p*_0*U*_] = [0.05, 0.13], these same designs exhibit substantial Type I error inflation, with the Type I error rate at the upper bound exceeding 0.25. In contrast, the BUDS-Worst-Regret objective selects a design that accounts for this uncertainty, yielding a maximum sample size of 89 and an average *EN* of 41.5. The Type I error rate remains below the planning *α* across the entire benchmark range, with a maximum value of 0.093.

A similar pattern is observed for TTE endpoints. Consider the Phase II trial of buparlisib using PFS6, with a planning benchmark of *S*_0_(*x*_0_) = 0.15 and a target survival probability of *S*_1_(*x*_0_) = 0.32 at *x*_0_ = 6 months, assuming an accrual rate of 18 patients per year (*α* = 0.10; *β* = 0.10). Under a single benchmark specification, the r-KJ design yields a maximum sample size of 31 while achieving nominal Type I error control at the planning value. However, when the plausible benchmark range [*S*_0*L*_(*x*_0_), *S*_0*U*_(x_0_)] = [0.09, 0.16] is considered, the Type I error increases to 0.13 at the upper bound. The BUDS-Worst-Regret-selected design requires only an additional six patients to control the Type I error across this range.

Together, these examples show that even modest levels of benchmark uncertainty can lead to materially different design choices and operating characteristics, particularly with respect to Type I error behavior. Details of the accompanying Shiny application, including example workflows and interface screenshots, are provided in the Supplementary Material 4.

## 4. Discussion

This work introduces the Benchmark Uncertainty Design Selection (BUDS) framework for two-stage, single-arm designs to address a common challenge in Phase II trial planning: uncertainty in the historical control benchmark. BUDS addresses this vulnerability by incorporating a clinically plausible range of benchmark values into the design-selection objective functions, while preserving the two-stage structure and error constraints.

The practical importance of benchmark uncertainty is most apparent during trial planning, when a historical control benchmark must be specified. When available evidence is heterogeneous, investigators typically select a representative value from the plausible range and apply a classic two-stage design. Calibration at a higher value provides greater protection against Type I error inflation if the true benchmark is higher than anticipated. However, any single-value approach remains vulnerable to benchmark misspecification. The BUDS framework eliminates the need to anchor planning on a single, potentially unrepresentative benchmark by making robustness an explicit planning criterion rather than a *post hoc* consideration. Specification of a benchmark range [*θ*_0*L*_, *θ*_0*U*_] promotes transparent discussion of clinically plausible benchmark values during protocol development and supports explicit, defensible design choices grounded in the available evidence.

Importantly, additional enrollment is not an intrinsic penalty of BUDS. It arises primarily because protecting against a higher plausible benchmark while maintaining power at a fixed alternative requires more information. The BUDS objectives determine how efficiency is distributed across the benchmark range among designs satisfying these constraints. A false positive from a non-robust Phase II trial can trigger a costly failure in Phase III, exposing a large patient population to a futile therapy (36, 37). The additional enrollment required by BUDS-chosen designs is therefore a rational strategy to mitigate this downstream risk. Efficiency can be moderated by narrowing the benchmark range, modestly lowering target power, or, when enrollment is capped, maximizing attainable power while preserving Type I error control across the range.

The proposed BUDS framework addresses uncertainty in the prespecified historical benchmark but does not eliminate the broader limitations inherent to single-arm studies. Although single-arm and randomized designs are not directly interchangeable, an important consideration is whether a randomized design becomes preferable when the required sample size for a single-arm study becomes large. At the same maximum sample size, a randomized design may have lower power than a comparable single-arm design (data not presented here), but it reduces reliance on historical benchmarks and provides better protection against bias from population differences, changes in clinical practice, and other sources of confounding. However, randomized trials may be infeasible because of limited patient populations, ethical or logistical constraints, increased cost, or the lack of an appropriate concurrent control. When the required sample size for a single-arm study approaches that of a feasible randomized trial, the improved internal validity afforded by randomization should be weighed carefully against the statistical and operational efficiency of the single-arm designs.

Currently, BUDS addresses uncertainty in the historical benchmark while treating the target alternative as prespecified. In practice, uncertainty may also exist regarding the magnitude of the clinically meaningful treatment effect. To examine the sensitivity of the BUDS-Worst-Regret objective to this assumption, we conducted sensitivity analyses across a range of alternative response rates for representative binary settings. As expected, larger treatment effects resulted in smaller sample sizes, whereas treatment effects closer to the upper plausible benchmark required substantially larger trials (Table S3 in Supplementary Material 3).

Although illustrated in oncology settings, BUDS applies broadly to single-arm Phase II studies in which the planning benchmark is uncertain. Future work will extend BUDS to jointly account for uncertainty in both historical benchmark and target efficacy rates. Other potential extensions include designs with early stopping for efficacy, randomized Phase II settings, and Bayesian analogues based on benchmark range posterior probabilities. For TTE outcomes, an important direction is the integration of BUDS principles with alternative modeling approaches.

## 5. Conclusion

BUDS offers a transparent and practical framework for incorporating benchmark uncertainty into trial design while maintaining Type I error control across the specified benchmark range and preserving the simplicity of the classic two-stage designs. The accompanying Shiny application provides a user-friendly platform for early phase trial planning and facilitates structured go/no-go decision-making.

## Data Availability

This manuscript presents a statistical methodology whose operating characteristics are assessed through exact computation across a range of prespecified planning scenarios; no empirical patient data were used or generated. All designs are fully reproducible using the open-source BUDS R package, available at: https://github.com/Ye-Lab-UMiami/BUDS. An interactive Shiny application for design implementation is also provided.

## Declarations

### Ethics approval and consent to participate

Not applicable.

### Consent for publication

Not applicable.

### Availability of data and materials

This manuscript presents a statistical methodology whose operating characteristics are assessed through exact computation across a range of prespecified planning scenarios; no empirical patient data were used or generated. All designs are fully reproducible using the open-source **BUDS** R package, available at: https://github.com/Ye-Lab-UMiami/BUDS. An interactive Shiny application for design implementation is also provided.

### Author’s contributions

All authors have been involved in the conception, design, analysis, and interpretation of this work and in drafting the manuscript. All authors read and approved the final manuscript.

## Acknowledgements

The authors acknowledge the support of the Biostatistics and Bioinformatics Shared Resource (BBSR) at the Sylvester Comprehensive Cancer Center, University of Miami.

## Abbreviations

BUDS: Benchmark Uncertainty Design Selection
TTE: time-to-event
KJ: Kwak and Jung
TMZ: temozolomide
PFS6: 6-month progression-free survival
EN: expected sample size
PET: probability of early termination
HR: hazard ratio
r-KJ: restricted-Kwak and Jung

