## Supplementary Material 1 for "BUDS: Benchmark Uncertainty Design Selection for Two-Stage Single-Arm Phase II Trials"

### **Mathematical Proofs**

In the BUDS framework, verifying Type I error control appears computationally daunting, as it requires checking the rejection probability at an infinite number of points within the benchmark range. However, two fundamental monotonicity properties of the binomial distribution render this problem tractable. This appendix provides formal statements and proofs of these properties, along with results on early termination, the structure of feasible sets, the well-definedness of BUDS objectives, and the existence and minimum sample size of feasible designs.

**Lemma 1 (Monotonicity of Rejection Probability):** For any fixed two-stage design (n_1_,r_1_;n,r), the probability of a "Go" decision, $\Pr_{p} \left( Go \right)$, is a non-decreasing function of the true response rate $p\in\left[ 0,1 \right]$.

Proof: Denote $\alpha\left( p \right)=\Pr_{p} (Go)$ for simplicity. Let $n_{2}=n-n_{1}$. We make the “Go” decision if and only if the following two conditions hold simultaneously: (1). $x_{1}>r\_1$ (2). $x_{1}+x_{2}>r$. Therefore,

$$\alpha\left( p \right)= \sum_{x_{1}=r_{1}+1}^{n_{1}} \binom{n_{1}}{x_{1}}p^{x_{1}}{(1-p)}^{n_{1}-x_{1}} \sum_{x_{2}=max(0, r-x_{1}+1)}^{n_{2}} \binom{n_{2}}{x_{2}}p^{x_{2}}\left( 1-p \right)^{n_{2}-x_{2}}$$

Denote $B\left( k;m, p \right)=\Pr\left( Bin\left( m, p \right)\leq k \right)=\sum_{i=0}^{k} \binom{m}{i}p^{i}\left( 1-p \right)^{m-i}$ the CDF of binomial distribution. We can write $\alpha(p)$ as

$$\alpha\left( p \right)= \sum_{x_{1}=r_{1}+1}^{n_{1}} \binom{n_{1}}{x_{1}}p^{x_{1}}{(1-p)}^{n_{1}-x_{1}}[1-B(r-x_{1};n_{2} , p)]$$

Claim 1: For $0<k<m$, $B\left( k;m, p \right)$ is strictly decreasing in $p$. It is called the probability of early termination (PET).

This claim can be proven by writing

$$\frac{d}{dp}B\left( k;m, p \right)= \sum_{i=0}^{k} \binom{m}{i}\left[ ip^{i-1}-\left( m-i \right)\left( 1-p \right)^{m-i-1} \right].$$

Note that

$$\binom{m}{i}i=m\binom{m-1}{i-1}, \binom{m}{i}\left( m-i \right)=m\binom{m-1}{i}$$

We have

$$\frac{d}{dp}B\left( k;m, p \right)= m\sum_{j=0}^{k-1} \binom{m-1}{j}p^{j}\left( 1-p \right)^{\left( m-1 \right)-j}-m\sum_{j=0}^{k} \binom{m-1}{j}p^{j}\left( 1-p \right)^{\left( m-1 \right)-j}=-m{\binom{m-1}{k}p}^{k}\left( 1-p \right)^{\left( m-1 \right)-k}<0,$$

and we have $B\left( k;m, p \right)$ strictly decreasing in $p$.

Let $X_{1}\sim Bin(n_{1}, p)$, $X_{2}\sim Bin(n_{2},p)$ and $X_{1}, X_{2}$ be independent. Note $\alpha\left( p \right)=\Pr\left( X_{1}>r_{1}, X_{1}+X_{2}>r \right)$. Define the set

$$A=\{\left( x_{1}, x_{2} \right):x_{1}>r_{1}, x_{1}+x_{2}>r\}$$

The set $A$ is increasing as for $\left( x_{1}^{'},x_{2}^{'} \right)\geq(x_{1}, x_{2})$, if $\left( x_{1},x_{2} \right)\in A$, then $\left( x_{1}^{'},x_{2}^{'} \right)\in A$.

For $U_{1},U_{2},\cdots,U_{n}\sim Unif(0,1)$ and independent of each other, define

$$I_{i}\left( p \right)= \boldsymbol{1}\left\{ U_{i}\leq p \right\},$$

Then for any $i$, $I_{i}\left( p \right)\sim Bernoulli(p)$, and $I_{i}\left( p_{2} \right)\geq I_{i}(p_{1})$ pointwisely for $p_{2}>p_{1}$. Let

$$X_{1}\left( p \right)=\sum_{i=1}^{n_{1}} I_{i}(p), X_{2}\left( p \right)= \sum_{i=n_{1}+1}^{n} I_{i}(p),$$

Then $X_{1}$ and $X_{2}$ are independent binomial distributed, and for $p_{2}>p_{1}$, $X_{1}\left( p_{2} \right)\geq X_{1}\left( p_{1} \right), X_{2}\left( p_{2} \right)\geq X_{2}(p_{1})$ pointwisely. Recall $A$ is increasing, we have

$$\boldsymbol{1}\left\{ \left( X_{1}\left( p_{1} \right),X_{2}\left( p_{1} \right) \right)\in A \right\}\leq\boldsymbol{1}\left\{ \left( X_{1}\left( p_{2} \right),X_{2}\left( p_{2} \right) \right)\in A \right\}$$

Take expectations on both sides, we have

$$\Pr\left( \left( X_{1}\left( p_{1} \right),X_{2}\left( p_{1} \right) \right)\in A \right)\leq\Pr\left( \left( X_{1}\left( p_{2} \right),X_{2}\left( p_{2} \right) \right)\in A \right)$$

i.e. $\alpha\left( p_{2} \right)\geq\alpha(p_{1})$ for $p_{2}>p_{1}$. $∎$

**Corollary 1 (Worst-Case Location for Type I Error):** As a direct consequence of Lemma 1, the supremum of the rejection probability over the benchmark range occurs at its upper boundary.

$$\sup_{p\in\left[ p_{0L},p_{0U} \right]} \Pr_{p} \left( \text{Go} \right)=\Pr_{p_{0U}} \left( \text{Go} \right)$$

This corollary is of immense practical importance. It reduces the complex task of controlling the Type I error over the entire interval to a simple check at a single point, p_0U_. The strong control constraint is satisfied if and only if $\Pr_{p_{0U}} \left( Go \right)\leq\alpha$.

A similar property holds for the expected sample size:

**Lemma 2 (Monotonicity of Expected Sample Size):** For any fixed design (n_1_,r_1_;n,r), the expected sample size, EN(p), is a non-decreasing function of p.

Proof: we have

$$EN\left( p \right)=n_{1}\Pr_{p} \left( X_{1}\leq r_{1} \right)+n\Pr_{p} \left( X_{1}>r_{1} \right)=n_{1}+n_{2}\Pr_{p} (X_{1}>r_{1})=n_{1}+n_{2}[1-B(r_{1};n_{1},p)]$$

When $r_{1}=n_{1}$, $B\left( n_{1};n_{1},p \right)=1$ for all $p$, and $EN\left( p \right)=n$ is a constant.

When $0\leq r_{1}<n_{1}$, we showed $B\left( r_{1};n_{1}, p \right)$ is strictly decreasing in $p$ in Lemma 1, so $1-B(r_{1};n_{1},p)$ is strictly increasing in $p$, i.e. $EN(p)$ is strictly increasing in $p$.

Therefore, $EN(p)$ is non-decreasing in $p$. $∎$

**Corollary 2 (Worst-Case Location for EN under H_0_):** The maximum expected sample size under the benchmark range occurs at p_0U_.

$$\sup_{p\in\left[ p_{0L},p_{0U} \right]} EN(p)=EN\left( p_{0U} \right)$$

Together, these properties of the binomial distribution make BUDS-selected design computation far more efficient – what would otherwise require exhaustive evaluation of Type I error across the entire benchmark range p ∈ [p_0L_, p_0U_] reduces to a single check at the upper boundary, p_0U._

**Remark:** The lemmas and corollaries above reveal that all three worst case operating characteristics ($\alpha\left( p \right), EN\left( p \right), PET$) under the benchmark range occur at the same point $p_{0U}$. This means that at the least favorable benchmark configuration, the design is most likely to produce a false positive and least likely to self-correct through early stopping.

**Proposition 1 (Nesting of Feasible Sets):** Let

$$D\left( \left[ a,b \right] \right)=D\left( \alpha,1-\beta,\left[ a,b \right],p_{1} \right)=\{d=\left( n_{1},r_{1};n,r \right):\Pr_{p_{1}} \left( Go \right)\geq1-\beta and\Pr_{b} \left( Go \right)\leq\alpha\}$$

denote the set of all two-stage designs satisfying the BUDS constraints for benchmark range $[a,b]$. If $\left[ a,b \right]\subseteq[c,d]$, then

$$D\left( \alpha,1-\beta,\left[ c,d \right],p_{1} \right)\subseteq D\left( \alpha,1-\beta,\left[ a,b \right],p_{1} \right)$$

Proof: Let  $\tilde{d}$ be any design in $D\left( \alpha,1-\beta,\left[ c,d \right],p_{1} \right)$. The power constraint depends only on $p_{1}$ and is independent of the benchmark range, so ${\Pr_{p_{1}}}^{\tilde{d}} \left( Go \right)\geq1-\beta$ is automatically satisfied for $[a,b]$. From Corollary 1, we have ${\Pr_{b}}^{\tilde{d}} \left( Go \right)\leq{\Pr_{d}}^{\tilde{d}} \left( Go \right)\leq\alpha$. Therefore, $\tilde{d}\in D\left( \alpha,1-\beta,\left[ a,b \right],p_{1} \right)$. $∎$

**Corollary 3 (Monotonicity of minimum sample size):** For any design characteristic $\phi(d)$:

$$\min_{d\in D([c,d])} \phi(d)\geq\min_{d\in D([a,b])} \phi(d)$$

Whenever $\left[ a,b \right]\subseteq[c,d]$, provided the feasible sets are non-empty.

This is a direct result from Proposition 1 and formalizes the empirical observation in Section 3.2 that the ‘price’ of robustness increases monotonically with interval width.

**Proposition 2 (Boundness of regret and well-definedness of the BUDS worst-regret objective):** For any feasible BUDS design $d\in D$ and any $p\in[p_{0L},p_{0U}]$:

a) $0\leq Regret_{d}\left( p \right)\leq n(d)$, where $n(d)$ is the maximum sample size of design $d$.
b) $sup_{p\in[p_{0L},p_{0U}]} Regret_{d}(p)$ exists and is finite.
c) $min_{d\in D}sup_{p\in[p_{0L},p_{0U}]} Regret_{d}(p)$ exists and is finite, provided $D\neq\emptyset$.

Proof:

a) By definition, $d^{*}\left( p \right)$ minimizes $EN\left( p \right)$ over all feasible designs. Therefore, $Regret_{d}\left( p \right)=EN_{d}\left( p \right)-EN_{d^{*}}\left( p \right)\geq0$, i.e. the lower bound exists.

The expected sample size of any design is bounded by its maximum sample size, i.e. $EN_{d}\left( p \right)\leq n(d)$ for all $p$. The expected sample size is also bounded by its stage-1 size and therefore bounded by 1. Thus,

$$Regret_{d}\left( p \right)=EN_{d}\left( p \right)-EN_{d^{*}}\left( p \right)\leq n\left( d \right)-1<n\left( d \right)<\infty$$

b) By part (a), $Regret_{d}\left( p \right)$ is a real-valued function on the compact interval $[p_{0L},p_{0u}]$ bounded above by $n\left( d \right)<\infty$. Every bounded set of real numbers has a finite supremum, therefore $sup_{p\in[p_{0L},p_{0U}]} Regret_{d}(p)$ exists and satisfies

$$0\leq sup_{p\in\left[ p_{0L},p_{0U} \right]} Regret_{d}\left( p \right)\leq n\left( d \right)$$

c) The feasible set $D$ is a finite set, and the minimum of finitely many finite real numbers exists and is finite. $∎$

**Proposition 3 (Existence of Feasible BUDS-selected designs):** For any $\left( \alpha,1-\beta,\left[ p_{0L},p_{0U} \right],p_{1} \right)$ with $0<\alpha<1$, $0<\beta<1$ and $p_{0U}<p_{1}$, there exists a finite integer $n^{*}$ such that for all $n_{max}\geq n^{*}$, the BUDS feasible set $D\left( \alpha,1-\beta,\left[ p_{0L},p_{0U} \right],p_{1} \right)$ is non-empty.

Proof: It suffices to exhibit a single feasible design for sufficiently large $n$. Consider a single-stage design as a special case of two-stage design $n_{1}=n$ and $r_{1}=n$, the design is reduced to enroll $n$ patients, declare “Go” if $X>r$, where $X\sim Bin(n,p)$. The two BUDS constraints become:

- Type I error: $\Pr_{p_{0u}} (X>r)=1-B\left( r;n,p_{0U} \right)\leq\alpha$
- Power: $\Pr_{p_{1}} (X>r)=1-B\left( r;n,p_{1} \right)\geq1-\beta$

These are equivalent to requiring an $r$ such that

$$B\left( r;n,p_{0U} \right)\geq1-\alpha and B\left( r;n,p_{1} \right)\leq\beta.$$

By the law of large numbers, $\frac{X}{n}\to p$ in probability, so under $p_{0u}$, the distribution of $X$ concentrates near $np_{0U}$, and under $p_{1}$ it concentrates near $np_{1}$. Any $r$ satisfying $np_{0U}<r< np_{1}$ will eventually separate the two distributions as $n\to\infty$. Take

$$r\left( n \right)=\left\lfloor n\bar{p} \right\rfloor, \bar{p}=\frac{p_{0U}+p_{1}}{2}.$$

Let $\Delta=p_{1}-p_{0U}>0$. Under $p=p_{0U}$, the standardized distance from $r$ to the mean is

$$\frac{r-np_{0U}}{\sqrt{np_{0U}\left( 1-p_{0U} \right)}}\approx\frac{n\left( \bar{p}-p_{0U} \right)}{\sqrt{np_{0U}\left( 1-p_{0U} \right)}}=\frac{\frac{\sqrt{n}\Delta}{2}}{\sqrt{p_{0U}\left( 1-p_{0U} \right)}}\to+\infty as n\to\infty,$$

which leads to $\Pr_{p_{0U}} (X>r)\to0<\alpha$. Similarly, under $p=p_{1}$,

$$\frac{r-np_{1}}{\sqrt{np_{1}\left( 1-p_{1} \right)}}=\frac{-\frac{\sqrt{n}\Delta}{2}}{\sqrt{p_{1}\left( 1-p_{1} \right)}}\to-\infty as n\to\infty,$$

which leads to $\Pr_{p_{1}} (X>r)\to1>1-\beta$.

Since both constraints hold and the due to continuity of probabilities, there exists a finite $n^{*}$ such that both constraints hold for all $n\geq n^{*}$. Because this single-stage design is a special case of a two-stage design, it belongs to the BUDS design space, and the feasible set is non-empty for all $n_{max}\geq n^{*}$. $∎$

**Corollary 4 (Minimum sample size growth rate):** Under the conditions of Proposition 3, the minimum sample size $n^{*}$ required satisfies

$$n^{*}\geq\frac{\left( z_{\alpha}\sqrt{p_{0U}(1-p_{0U})}+z_{\beta}\sqrt{p_{1}(1-p_{1})} \right)^{2}}{\left( p_{1}-p_{0U} \right)^{2}}$$

where $z_{\alpha}=\Phi^{-1}(1-\alpha)$ and $z_{\beta}=\Phi^{-1}(1-\beta)$, and $\Phi(\cdot)$ is the CDF of standard normal distribution. In particular,

$$n^{*}=\Omega\left( \frac{1}{\left( p_{1}-p_{0U} \right)^{2}} \right)$$

As $p_{0U}\to p_{1}$, $n^{*}\to\infty$.

Proof: We derive a necessary condition on $n$ for any $r$ to satisfy both Type I error and power constraints simultaneously. Under the normal assumption, we need

- Type I error: $r\geq np_{0U}+z_{\alpha}\sqrt{np_{0U}(1-p_{0U})}$
- Power: $r\leq np_{1}- z_{\beta}\sqrt{{np}_{1}\left( 1-p_{1} \right)}$

For any valid $r$, we must have

$$np_{1}- z_{\beta}\sqrt{{np}_{1}\left( 1-p_{1} \right)}\geq np_{0U}+z_{\alpha}\sqrt{{np}_{0U}\left( 1-p_{0U} \right)}$$

Rearrange and divide both sides by $\sqrt{n}>0$, we have

$$\sqrt{n}\left( p_{1}-p_{0U} \right)\geq z_{\alpha}\sqrt{p_{0U}\left( 1-p_{0U} \right)}+z_{\beta}\sqrt{p_{1}\left( 1-p_{1} \right)}$$

Rearrange again and square both sides, we have

$$n^{*}\geq\frac{\left( z_{\alpha}\sqrt{p_{0U}(1-p_{0U})}+z_{\beta}\sqrt{p_{1}(1-p_{1})} \right)^{2}}{\left( p_{1}-p_{0U} \right)^{2}}$$

This implies there’s no single-stage design (and hence no two-stage design) can be feasible with less than the above number of patients.

For the asymptotic rate, note that for fixed $\alpha, \beta, p_{1}$ and $p_{0U}\to p_{1}$, $z_{\alpha}\sqrt{p_{0U}(1-p_{0U})}+z_{\beta}\sqrt{p_{1}(1-p_{1})}\to(z_{\alpha}+z_{\beta})\sqrt{p_{1}(1-p_{1})}$, and hence

$$n^{*}\geq\frac{\left( z_{\alpha}+z_{\beta} \right)^{2}p_{1}\left( 1-p_{1} \right)}{\left( p_{1}-p_{0U} \right)^{2}}\left( 1+o\left( 1 \right) \right)\to\infty,$$

And this conforms

$$n^{*}=\Omega\left( \frac{1}{\left( p_{1}-p_{0U} \right)^{2}} \right)$$

$∎$
