## Supplementary Material 2 for "BUDS: Benchmark Uncertainty Design Selection for Two-Stage Single-Arm Phase II Trials"

**Restricted-Kwak and Jung BUDS theorem**

**Notation recap**

We borrow the notations from Belin et. al. (1). Consider a one-sample setting in which patients are monitored for a time-to-event endpoint with hazard rate $\lambda$. For each patient $i$, let $A_{i}, i=1,2,\cdots,n_{k}$ denote the accrual time for each patient; $T_{i}$ the failure time; $DA_{k}$ be the calendar time of stage $k$ analysis; $C_{i}=DA_{k}-A_{i}$ be the administrative censoring time; $x_{0}$ be the restricted follow-up time; and $X_{i}=\min\left( T_{i},x_{0},DA_{k}-A_{i} \right)$ be the observed time with event indicator $\delta_{i}=1\{X_{i}=t\}$. Let $F\left( t \right)$ denote the survival function of $T_{i}$, and $\Lambda\left( t \right)=\int_{0}^{t} \lambda\left( u \right)du$ the cumulative hazard. The event counting process and at-risk indicator are $N_{i}\left( t,DA_{k} \right)=1\{X_{i}\leq t,\delta_{i}=1\}$ and $Y_{i}(t,DA_{k})=1\{X_{i}\geq t\}$ respectively.

We denote $n_{1}$ as the number of patients enrolled in stage 1, $n_{2}$ the total number of patients enrolled by the end of stage 2, $t_{a}$ the accrual period where patients are enrolled over $[0,t_{a}]$. Under uniform accrual, the survival function of $C_{i}$ is $G\left( t,DA_{k} \right)=min\{1, \left( DA_{k}-t \right)_{+}/t_{a}\}$ for $0\leq t\leq x_{0}$. Note $G\left( t,DA_{2} \right)=1$ for all $t\leq x_{0}$ since $DA_{2}=t_{a}+x_{0}.$

We assume exponential survival, i.e. the survival function is $S\left( t \right)=e^{-\lambda t}$, and uniform accrual over $[0,t_{a}]$. We consider a hypothesis test:

$$H_{0}:\lambda\geq\lambda_{0L} v.s. H_{1}: \lambda<\lambda_{0L}$$

Since every choice in the benchmark range is feasible, we choose a design point $\lambda_{d}\in\left[ \lambda_{0L},\lambda_{0U} \right]$, and a design $\mathcal{D}$ is calibrated at this point if futility boundaries $c_{1}$ and $c_{2}$ are chosen so that type I error equals $\alpha$ when the true hazard rate is exactly $\lambda_{d}$, and the power equals $1-\beta$ at a chosen $\lambda_{1}<\lambda_{0L}$. Note that after we find $\mathcal{D}$, the type I error will be evaluated at evaluation point $\lambda_{0}\in[\lambda_{0L},\lambda_{0U}]$.

**Single stage design**

Let $W$ be the counting process of the one-sample log-rank test at calendar time of analysis $DA$:

$$W=\frac{1}{\sqrt{n}}\sum_{i=1}^{n} \int_{0}^{\infty} \left[ dN_{i}\left( t,DA \right)-Y_{i}\left( t,DA \right)d\Lambda_{d}\left( t \right) \right].$$

Under $H_{0}$, for any calibration $\lambda_{0}$ (denoted as $\lambda_{d}$ in this context for convenience) we pick, $W$ is approximately normal with mean 0 and variance can be estimated by

$$\hat{\sigma}_{0,d}^{2}=\frac{1}{n}\sum_{i=1}^{n} \int_{0}^{\infty} Y_{i}\left( t,DA \right)d\Lambda_{d}(t)$$

We reject $H_{0}$ under significant level $\alpha$ if $z=\frac{W}{\hat{\sigma}_{0,d}}< -z_{1-\alpha}$.

Under the evaluation point $\lambda_{0}$ which also stays in $[\lambda_{0L},\lambda_{0U}]$ but might differ from $\lambda_{d}$, we have the following uniform convergence in probability:

$$\frac{1}{n}\sum_{i=1}^{n} Y_{i}\left( t,DA \right)\underset{\to}{u.p.}S_{0}\left( t \right)G\left( t,DA \right)1\left\{ x_{0}\geq t \right\},$$

Where $S_{0}(t)$ is the survival function with $\lambda=\lambda_{0}$. Hence, we can write

$$\hat{\sigma}_{0,d}^{2}\to\sigma_{0}^{2}\left( \lambda_{0},\lambda_{d} \right)=\int_{0}^{x_{0}} S_{0}\left( t \right)G\left( t,DA \right)d\Lambda(t)=\int_{0}^{x_{0}} e^{-\lambda_{0}t}G\left( t,DA \right)\lambda_{d}dt=\lambda_{d}\int_{0}^{x_{0}} e^{-\lambda_{0}t}G\left( t,DA \right)dt.$$

Under $\lambda_{0}$, we shift the counting process $W$, and now it’s approximately Gaussian with mean $\sqrt{n}\omega$, where

$$\omega=\omega\left( \lambda_{0},\lambda_{d} \right)=\int_{0}^{x_{0}} S_{0}\left( t \right)G\left( t,DA \right)d\left( \Lambda_{0}\left( t \right)-\Lambda_{d}\left( t \right) \right)=(\lambda_{0}-\lambda_{d})\int_{0}^{x_{0}} e^{-\lambda_{0}t}G(t,DA)dt$$

And the variance of $W$ can be approximated by

$${\sigma'}_{0}^{2}\left( \lambda_{0} \right)=\int_{0}^{\infty} G\left( t,DA \right)S_{0}\left( t \right)d\Lambda_{0}(t)=\lambda_{0}\int_{0}^{\infty} e^{-\lambda_{0}t}G\left( t,DA \right)dt.$$

And the type I error is

$$\alpha\left( \lambda_{0} \right)=P_{\lambda_{0}}\left( \frac{W}{\hat{\sigma}_{0,d}}<-z_{1-\alpha} \right)=\Phi\left( \frac{-\sigma_{0}\left( \lambda_{0},\lambda_{d} \right)z_{1-\alpha}-\sqrt{n}\omega\left( \lambda_{0},\lambda_{d} \right)}{{\sigma'}_{0}\left( \lambda_{0} \right)} \right).$$

To evaluate power, we let $\tilde{\lambda}=(\lambda_{1}+\lambda_{d})/2$ and $\tilde{S}\left( t \right)=e^{-\tilde{\lambda}t}$, the variance is

$$\tilde{\sigma}_{1}^{2}=\tilde{\lambda}\int_{0}^{x_{0}} e^{-\tilde{\lambda}t}G(t,DA)dt,$$

And the power function is defined by

$$1-\beta=P(\frac{W}{\tilde{\sigma}_{1}}<-z_{1-\alpha}|H_{1})=P((W-\sqrt{n}\omega(\lambda_{1},\lambda_{d}))/\tilde{\sigma}_{1}<(-\sigma_{0}\left( \lambda_{1},\lambda_{d} \right)z_{1-\alpha}-\sqrt{n}\omega\left( \lambda_{1},\lambda_{d} \right))/\tilde{\sigma}_{1})$$

Note: if we have single benchmark value $\lambda=\lambda_{d}$, $\alpha\left( \lambda_{d} \right)=\Phi\left( -z_{1-\alpha} \right)=\alpha$

**Two-Stage design**

We consider two counting processes $W_{1}$ and $W_{2}$, where

$$W_{k}=\frac{1}{\sqrt{n_{k}}}\sum_{i=1}^{n} \int_{0}^{\infty} \left[ dN_{i}\left( t,DA_{k} \right)-Y_{i}\left( t,DA_{k} \right)d\Lambda_{d}\left( t \right) \right], k=1,2.$$

Similar to single stage analysis, the variance of $W_{k}$ can be approximated by

$$\hat{\sigma}_{0k}^{2}=\frac{1}{n_{k}}\sum_{i=1}^{n_{k}} \int Y_{i}\left( t,DA_{k} \right)d\Lambda_{d}\left( t \right)\to\lambda_{d}\int_{0}^{x_{0}} e^{-\lambda_{0}t}G(t,DA_{k})dt=\sigma_{0k}^{2}(\lambda_{0},\lambda_{d})$$

Plug in the exact formula of $G(t,DA_{k})$, we have

$$\sigma_{01}^{2}\left( \lambda_{0},\lambda_{d} \right)=\left\{ \begin{aligned} \frac{\lambda_{d}}{\lambda_{0}t_{a}}\left[ DA_{1}+\frac{e^{-\lambda_{0}DA_{1}}}{\lambda_{0}}-\frac{1}{\lambda_{0}} \right], DA_{1}<x_{0} \\ \frac{\lambda_{d}}{\lambda_{0}t_{a}}\left[ DA_{1}\left( 1-e^{-\lambda_{0}x_{0}} \right)+x_{0}e^{-\lambda_{0}x_{0}}+\frac{e^{-\lambda_{0}x_{0}}}{\lambda_{0}}-\frac{1}{\lambda_{0}} \right],DA_{1}\geq x_{0} \end{aligned} \right.$$

And

$$\sigma_{02}^{2}\left( \lambda_{0},\lambda_{d} \right)=\frac{\lambda_{d}}{\lambda_{0}}\left( 1-e^{-\lambda_{0}x_{0}} \right)$$

Analogous to single-stage analysis, the mean under evaluation point $\lambda_{0}$ is $E_{\lambda_{k}}\left( W_{k} \right)=\sqrt{n_{k}}\omega_{k}(\lambda_{0},\lambda_{d})$, where under exponential survival and uniform accrual assumptions:

$$\omega_{1}\left( \lambda_{0},\lambda_{d} \right)=\left\{ \begin{aligned} \frac{\lambda_{0}-\lambda_{d}}{\lambda_{e}t_{a}}\left[ DA_{1}+\frac{e^{-\lambda_{0}DA_{1}}}{\lambda_{0}}-\frac{1}{\lambda_{0}} \right], DA_{1}<x_{0} \\ \frac{\lambda_{0}-\lambda_{d}}{\lambda_{0}t_{a}}\left[ DA_{1}\left( 1-e^{-\lambda_{0}x_{0}} \right)+x_{0}e^{-\lambda_{0}x_{0}}+\frac{e^{-\lambda_{0}x_{0}}}{\lambda_{0}}-\frac{1}{\lambda_{0}} \right],DA_{1}\geq x_{0} \end{aligned} \right.$$

And

$$\omega_{2}\left( \lambda_{0},\lambda_{d} \right)=\left( 1-\frac{\lambda_{d}}{\lambda_{0}} \right)[1-e^{-\lambda_{0}x_{0}}]$$

For type I error evaluation, $\sigma_{0k}^{2}$ can be approximated as $\lambda_{0}\int_{0}^{x_{0}} e^{-\lambda_{0}t}G(t,DA_{k})dt$, and under the assumptions,

$${\sigma'}_{01}^{2}\left( \lambda_{0} \right)=\left\{ \begin{aligned} \frac{1}{t_{a}}\left[ DA_{1}+\frac{e^{-\lambda_{0}DA_{1}}}{\lambda_{0}}-\frac{1}{\lambda_{0}} \right], DA_{1}<x_{0} \\ \frac{1}{t_{a}}\left[ DA_{1}\left( 1-e^{-\lambda_{0}x_{0}} \right)+x_{0}e^{-\lambda_{0}x_{0}}+\frac{e^{-\lambda_{0}x_{0}}}{\lambda_{0}}-\frac{1}{\lambda_{0}} \right],DA_{1}\geq x_{0} \end{aligned} \right.$$

And

$${\sigma'}_{02}^{2}\left( \lambda_{0} \right)=1-e^{-\lambda_{0}x_{0}}$$

Therefore, the correlation coefficient between $W_{1}$ and $W_{2}$ under evaluation point $\lambda_{0}$ is

$$\rho_{H_{0}}\left( \lambda_{0} \right)=\frac{{\sigma'}_{01}(\lambda_{0})}{{\sigma'}_{02}(\lambda_{0})}$$

For power evaluation, we still define $\tilde{\lambda}=(\lambda_{1}+\lambda_{d})/2$, and

$$\tilde{\sigma}_{11}^{2}=\left\{ \begin{aligned} \frac{1}{t_{a}}\left[ DA_{1}+\frac{e^{-\tilde{\lambda}DA_{1}}}{\tilde{\lambda}}-\frac{1}{\tilde{\lambda}} \right], DA_{1}<x_{0} \\ \frac{1}{t_{a}}\left[ DA_{1}\left( 1-e^{-\tilde{\lambda}x_{0}} \right)+x_{0}e^{-\tilde{\lambda}x_{0}}+\frac{e^{-\tilde{\lambda}x_{0}}}{\tilde{\lambda}}-\frac{1}{\tilde{\lambda}} \right],DA_{1}\geq x_{0}, \end{aligned} \right.$$

$$\sigma_{12}^{2}=1-e^{-\tilde{\lambda}x_{0}}$$

The correlation is then

$$\rho_{H_{1}}=\frac{\tilde{\sigma}_{11}}{\tilde{\sigma}_{12}}$$

For design $\mathcal{D}$ with futility boundaries $c_{1},c_{2}$ calibrated at $\lambda_{d}$, the test statistic $Z_{k}$ is not approximately $N(0,1)$ when evaluation point $\lambda_{0}\neq\lambda_{d}$, but with expected value $\omega_{k}(\lambda_{0},\lambda_{d})$ and standard deviation $\sigma_{0k}(\lambda_{0})/\sigma_{0k}(\lambda_{0},\lambda_{d})$. Define

$$\bar{c}_{k}\left( \lambda_{0} \right)=\frac{\sigma_{0k}\left( \lambda_{0},\lambda_{d} \right)}{{\sigma'}_{0k}\left( \lambda_{0} \right)}\left( c_{k}-\frac{\omega_{k}\left( \lambda_{0},\lambda_{d} \right)\sqrt{n_{k}}}{\sigma_{0k}\left( \lambda_{0},\lambda_{d} \right)} \right)$$

Then the type I error of design $\mathcal{D}$ evaluated at $\lambda_{0}$ is

$$\alpha\left( \lambda_{0}\mathcal{;D} \right)=\Phi_{2}\left( \bar{c}_{1}\left( \lambda_{0} \right),\bar{c}_{2}\left( \lambda_{0} \right);\rho_{H_{0}}\left( \lambda_{0} \right) \right)=\int_{-\infty}^{\bar{c}_{2}\left( \lambda_{0} \right)} \phi\left( z \right)\Phi\left( \frac{\bar{c}_{1}\left( \lambda_{0} \right)-\rho_{H_{0}}\left( \lambda_{0} \right)z}{\sqrt{1-\rho_{H_{0}}\left( \lambda\right)^{2}}} \right)dz (1)$$

Similarly define

$$\bar{c}_{k}^{(1)}=\frac{\sigma_{0k}\left( \lambda_{1},\lambda_{d} \right)}{\tilde{\sigma}_{1k}}\left( c_{k}-\frac{\omega_{k}\left( \lambda_{1},\lambda_{d} \right)\sqrt{n_{k}}}{\sigma_{0k}\left( \lambda_{1},\lambda_{d} \right)} \right),$$

And the power equals to

$$\int_{-\infty}^{\bar{c}_{2}^{(1)}} \phi\left( z \right)\Phi\left( \frac{\bar{c}_{1}^{(1)}-\rho_{H_{1}}z}{\sqrt{1-\rho_{H_{1}}^{2}}} \right)dz$$

The PET under $\lambda_{0}$ is then

$$PET\left( \lambda_{0}\mathcal{;D} \right)=P_{\lambda_{0}}\left( Z_{1}>c_{1} \right)=1-\Phi(\bar{c}_{1}\left( \lambda_{0} \right))$$

And EN is

$$EN\left( \lambda_{0}\mathcal{;D} \right)=n_{1}+(1-PET(\lambda_{0}\mathcal{;D))(}n_{2}-n_{1})$$

**r-KJ-BUDS proof**

Recall we define a two-stage design $\mathcal{D}$ calibrated at a design point $\lambda_{d}\in[\lambda_{0L},\lambda_{0U}]$ with boundaries $c_{1},c_{2}\mathbb{\in R}$. We evaluate type I error at an evaluation point $\lambda_{0}\in[\lambda_{0L},\lambda_{0U}]$.

Assume exponential survival with hazard rate $\lambda$ and uniform accrual over $[0,t_{a}]$. For stage $k=1,2$, define

$$I_{k}\left( \lambda_{0} \right)=\int_{0}^{x_{0}} e^{-\lambda_{0}t}G(t,DA_{k})dt$$

Since $DA_{2}=t_{a}+x_{0}$, we have $G\left( t,DA_{2} \right)=1$ for $t\leq x_{0}$, therefore.

$$I_{2}\left( \lambda_{0} \right)=\int_{0}^{x_{0}} e^{-\lambda_{0}t}dt=\frac{1-e^{-\lambda x_{0}}}{\lambda}$$

From the formulas we had,

$$\sigma_{0k}^{2}\left( \lambda_{0},\lambda_{d} \right)=\lambda_{d}I_{k}\left( \lambda_{0} \right),{\sigma'}_{0k}^{2}\left( \lambda_{0} \right)=\lambda_{0}I_{k}\left( \lambda_{e} \right),\omega_{k}\left( \lambda_{0},\lambda_{d} \right)=\left( \lambda_{0}-\lambda_{d} \right)I_{k}\left( \lambda_{0} \right) (2)$$

Therefore,

$$\bar{c_{k}}\left( \lambda_{0} \right)=\frac{\sigma_{0k}\left( \lambda_{0},\lambda_{d} \right)c_{k}-\omega_{k}(\lambda_{0},\lambda_{d})\sqrt{n_{k}}}{{\sigma'}_{0k}(\lambda_{0})}$$

And the correlation is

$$\rho_{H_{0}}\left( \lambda_{0} \right)=\frac{{\sigma'}_{01}(\lambda_{0})}{{\sigma'}_{02}(\lambda_{0})}=\sqrt{\frac{I_{1}(\lambda_{0})}{I_{2}(\lambda_{0})}}$$

The type I error is

$$\alpha\left( \lambda_{0}\mathcal{,D} \right)=\Phi_{2}(\bar{c_{1}}\left( \lambda_{0} \right),\bar{c_{2}}\left( \lambda_{0} \right);\rho_{H_{0}}\left( \lambda_{0} \right))$$

Where $\Phi_{2}\left( a,b;\rho\right)=P(Z_{1}\leq a,Z_{2}\leq b)$ for a standard bivariate normal vector $(Z_{1},Z_{2})$ with correlation $\rho$.

Further, note

$$\frac{\sigma_{0k}(\lambda_{0},\lambda_{d})}{{\sigma'}_{0k}(\lambda_{0})}=\sqrt{\frac{\lambda_{d}}{\lambda_{0}}}$$

And

$$\frac{\omega_{k}(\lambda_{0},\lambda_{d})\sqrt{n_{k}}}{\sigma_{0k}(\lambda_{0})}=\frac{\left( \lambda_{0}-\lambda_{d} \right)I_{k}\left( \lambda_{0} \right)\sqrt{n_{k}}}{\sqrt{\lambda_{d}I_{k}(\lambda_{0})}}=\frac{(\lambda_{0}-\lambda_{d})\sqrt{n_{k}I_{k}(\lambda_{0})}}{\sqrt{\lambda_{d}}}$$

Hence

$$\bar{c_{k}}\left( \lambda_{0} \right)=\frac{\sqrt{\lambda_{d}}c_{k}-(\lambda_{0}-\lambda_{d})\sqrt{n_{k}I_{k}(\lambda_{0})}}{\sqrt{\lambda_{0}}}$$

Define $h_{k}\left( \lambda_{0} \right)=\sqrt{n_{k}I_{k}(\lambda_{0})}$ and $S_{k}\left( \lambda_{0} \right)=\sqrt{\lambda_{d}}c_{k}-\left( \lambda_{e}-\lambda_{d} \right)h_{k}(\lambda_{0})$, so we can write $\bar{c_{k}}\left( \lambda_{0} \right)$ as

$$\bar{c_{k}}\left( \lambda_{0} \right)=\frac{S_{k}\left( \lambda_{0} \right)}{\sqrt{\lambda_{0}}}$$

**Lemma 1.** For each $k=1,2$, the function $I_{k}(\lambda_{0})$ is strictly positive and strictly decreasing for $\lambda_{e}$.

Proof: From definition of $G\left( t,DA_{k} \right)$, it is non-negative and not identically zero on $[0,x_{0}]$, so

$I_{k}\left( \lambda_{0} \right)=\int_{0}^{x_{0}} e^{-\lambda_{0}t}G(t,DA_{k})dt>0$. Take derivative under the integral, we have

$I_{k}^{'}(\lambda_{0})=-\int_{0}^{x_{0}} {te}^{-\lambda_{0}t}G\left( t,DA_{k} \right)dt<0$.

**Proposition 1.** Let

$$q\left( \lambda_{0} \right)=\lambda_{0}+\lambda_{d}-\lambda_{0}\left( \lambda_{0}-\lambda_{d} \right)_{+}x_{0}$$

Assume for each $k=1,2$ and all $\lambda_{0}\in[\lambda_{0L},\lambda_{0U}]$, $q\left( \lambda_{0} \right)>0$, $h_{k}\left( \lambda_{0} \right)>\frac{\sqrt{\lambda_{d}}|c_{k}|}{q(\lambda_{0})}$. Then ${\bar{c_{k}}}^{'}\left( \lambda_{0} \right)<0$ on $[\lambda_{0L}, \lambda_{0U}]$ for $k=1,2$.

Proof: We have

$$h_{k}^{'}\left( \lambda_{0} \right)=\frac{n_{k}I_{k}^{'}(\lambda_{0})}{2\sqrt{n_{k}I_{k}(\lambda_{0})}}=\frac{I_{k}^{'}\left( \lambda_{0} \right)}{2I_{k}\left( \lambda_{0} \right)}h_{k}(\lambda_{0})$$

Define

$$m_{k}\left( \lambda_{0} \right)=\frac{\int_{0}^{x_{0}} te^{-\lambda_{0}t}G(t,DA_{k})dt}{\int_{0}^{x_{0}} e^{-\lambda_{0}t}G(t,DA_{k})dt}$$

Then $I_{k}^{'}\left( \lambda_{0} \right)=-m_{k}\left( \lambda_{0} \right)I_{k}(\lambda_{0})$, and

$$h_{k}^{'}\left( \lambda_{0} \right)=\frac{-m_{k}\left( \lambda_{0} \right)}{2}h_{k}(\lambda_{0})$$

Therefore,

$$F_{k}^{'}\left( \lambda_{0} \right)=-h_{k}\left( \lambda_{0} \right)-\left( \lambda_{0}-\lambda_{d} \right)h_{k}^{'}\left( \lambda_{0} \right)=-h_{k}\left( \lambda_{0} \right)+\frac{\lambda_{0}-\lambda_{d}}{2}m_{k}\left( \lambda_{0} \right)h_{k}\left( \lambda_{0} \right).$$

Since $\bar{c_{k}}\left( \lambda_{0} \right)=F_{k}\left( \lambda_{0} \right)\lambda^{-\frac{1}{2}}$,

$${\bar{c_{k}}}^{'}(\lambda_{0})=\frac{2\lambda_{0}F_{k}^{'}\left( \lambda_{0} \right)-F_{k}(\lambda_{0})}{2\lambda^{3/2}}$$

And the numerator is

$$2\lambda_{0}\left( -h_{k}\left( \lambda_{0} \right)+\frac{\lambda_{0}-\lambda_{d}}{2}m_{k}\left( \lambda_{0} \right)h_{k}\left( \lambda_{0} \right) \right)-F_{k}\left( \lambda_{0} \right)=h_{k}\left( \lambda_{0} \right)\left[ -\left( \lambda_{0}+\lambda_{d} \right)+\lambda_{0}\left( \lambda_{0}-\lambda_{d} \right)m_{k}\left( \lambda_{0} \right) \right]-\sqrt{\lambda_{d}}c_{k}\leq-q\left( \lambda_{0} \right)h_{k}\left( \lambda_{0} \right)-\sqrt{\lambda_{d}}c_{k}.$$

The last inequality comes from $0\leq m_{k}\left( \lambda_{0} \right)\leq x_{0}$.

If $c_{k}\geq0$, then $-q\left( \lambda_{0} \right)h_{k}\left( \lambda_{0} \right)-\sqrt{\lambda_{d}}c_{k}<0$ follows from $q\left( \lambda_{0} \right)>0$. If $c_{k}<0$, then $q\left( \lambda_{0} \right)h_{k}\left( \lambda_{0} \right)>\sqrt{\lambda_{d}}|c_{k}|$, and $-q\left( \lambda_{0} \right)h_{k}\left( \lambda_{0} \right)-\sqrt{\lambda_{d}}c_{k}<0$ still holds. Hence $2\lambda_{0}F_{k}^{'}\left( \lambda_{0} \right)-F_{k}\left( \lambda_{0} \right)<0$ and ${\bar{c_{k}}}^{'}\left( \lambda_{0} \right)<0$.

**Lemma 2.** $\rho_{H_{0}}\left( \lambda_{0} \right)$ is non-decreasing in $\lambda_{0}$ on $(0,\infty)$.

Proof: From

$$\rho_{H_{0}}\left( \lambda_{0} \right)=\frac{{\sigma'}_{01}(\lambda_{0})}{{\sigma'}_{02}(\lambda_{0})}=\sqrt{\frac{I_{1}(\lambda_{0})}{I_{2}(\lambda_{0})}},$$

We have

$$\rho_{H_{0}}^{'}\left( \lambda_{0} \right)=\frac{\rho_{H_{0}}\left( \lambda_{0} \right)}{2}\left( \frac{I_{1}^{'}\left( \lambda_{0} \right)}{I_{1}\left( \lambda_{0} \right)}-\frac{I_{2}^{'}\left( \lambda_{0} \right)}{I_{2}\left( \lambda_{0} \right)} \right)=\frac{\rho_{H_{0}}\left( \lambda_{0} \right)}{2}(m_{2}\left( \lambda_{0} \right)-m_{1}(\lambda_{0}))$$

Since $G\left( t,DA_{2} \right)=1$ and $G\left( t,DA_{1} \right)=min\{1, \left( DA_{1}-t \right)_{+}/t_{a}\}$ is non-increasing in $t$ on $[0,x_{0}]$. From Chebyshev integral inequality, under the positive measure $d\nu\left( t \right)=e^{-\lambda_{0}t}dt$, we have

$$\left( \int_{0}^{x_{0}} tG(t,DA_{1})d\nu\left( t \right) \right)\left( \int_{0}^{x_{0}} d\nu\left( t \right) \right)\leq\left( \int_{0}^{x_{0}} td\nu\left( t \right) \right)\left( \int_{0}^{x_{0}} G(t,DA_{1})d\nu\left( t \right) \right).$$

Divide both sides by $\left( \int_{0}^{x_{0}} d\nu\left( t \right) \right)\left( \int_{0}^{x_{0}} G(t,DA_{1})d\nu\left( t \right) \right)$, since $\left( \int_{0}^{x_{0}} d\nu\left( t \right) \right)\left( \int_{0}^{x_{0}} G(t,DA_{1})d\nu\left( t \right) \right)$ is positive, we have

$$m_{1}\left( k \right)= \frac{\int_{0}^{x_{0}} te^{-\lambda_{0}t}G(t,DA_{1})dt}{\int_{0}^{x_{0}} e^{-\lambda_{0}t}G(t,DA_{1})dt}\leq\frac{\int_{0}^{x_{0}} te^{-\lambda_{0}t}dt}{\int_{0}^{x_{0}} e^{-\lambda_{0}t}dt}=m_{2}(k)$$

i.e. $\rho_{H_{0}}^{'}\left( \lambda_{0} \right)\geq0$, $\rho_{H_{0}}\left( \lambda_{0} \right)$ is non-decreasing in $\lambda_{0}$ on $(0,\infty)$.

For conveniency, write $s\left( \lambda_{0} \right)= \sqrt{1-\rho_{H_{0}}\left( \lambda_{0} \right)^{2}}$. For $Z_{1},Z_{2}\sim N\left( 0,1 \right)$ with correlation $\rho_{H_{0}}(\lambda_{0})$, we have

$$Z_{2}|Z_{1}=\bar{c_{1}}(\lambda_{0})\sim N\left( \rho_{H_{0}}\left( \lambda_{0} \right)\bar{c_{1}}(\lambda_{0}),s\left( \lambda_{0} \right)^{2} \right),$$

So the function $\Phi_{2}(a,b;\rho)$ satisfies:

$$\left. \frac{\partial\Phi_{2}}{\partial a}\left( a,b;\rho\right) \right|_{a=\bar{c_{1}}(\lambda_{0}),b=\bar{c_{2}}(\lambda_{0}),\rho=\rho_{H_{0}}(\lambda_{0})}=\phi\left( \bar{c_{1}}(\lambda_{0}) \right)\Phi\left( \frac{\bar{c_{2}}(\lambda_{0})-\bar{c_{1}}{(\lambda_{0})\rho}_{H_{0}}\left( \lambda_{0} \right)}{s\left( \lambda_{0} \right)} \right).$$

Similarly,

$$\left. \frac{\partial\Phi_{2}}{\partial b}\left( a,b;\rho\right) \right|_{a=\bar{c_{1}}(\lambda_{0}),b=\bar{c_{2}}(\lambda_{0}),\rho=\rho_{H_{0}}(\lambda_{0})}=\phi\left( \bar{c_{2}}(\lambda_{0}) \right)\Phi\left( \frac{\bar{c_{1}}(\lambda_{0})-\bar{c_{2}}{(\lambda_{0})\rho}_{H_{0}}\left( \lambda_{0} \right)}{s\left( \lambda_{0} \right)} \right).$$

Also,

$$\frac{\partial\Phi_{2}}{\partial\rho}\left( a,b;\rho\right)=\phi_{2}\left( a,b;\rho\right),$$

And all these three partial derivatives are non-negative.

**Theorem 1 (Monotonicity):** Assume $q\left( \lambda_{0} \right)>0$, and $h_{k}\left( \lambda_{0} \right)>\frac{\sqrt{\lambda_{d}}|c_{k}|}{q(\lambda_{0})}$, so that ${\bar{c_{k}}}^{'}\left( \lambda_{0} \right)<0$ on $[\lambda_{0L}, \lambda_{0U}]$ for $k=1,2$. If for every $\lambda_{0}\in[\lambda_{0L},\lambda_{0U}]$,

$$\phi\left( \bar{c_{1}}\left( \lambda_{0} \right) \right)\Phi\left( \frac{\bar{c_{2}}\left( \lambda_{0} \right)-\bar{c_{1}}{\left( \lambda_{0} \right)\rho}_{H_{0}}\left( \lambda_{0} \right)}{s\left( \lambda_{0} \right)} \right)\left| {\bar{c_{1}}}^{'}\left( \lambda_{0} \right) \right|+ \phi\left( \bar{c_{2}}\left( \lambda_{0} \right) \right)\Phi\left( \frac{\bar{c_{1}}\left( \lambda_{0} \right)-\bar{c_{2}}{\left( \lambda_{0} \right)\rho}_{H_{0}}\left( \lambda_{0} \right)}{s\left( \lambda_{0} \right)} \right)\left| {\bar{c_{2}}}^{'}\left( \lambda_{0} \right) \right|\geq\phi_{2}\left( \bar{c_{1}}\left( \lambda_{0} \right),\bar{c_{2}}\left( \lambda_{0} \right);\rho_{H_{0}}\left( \lambda_{0} \right) \right){\rho^{'}}_{H_{0}}\left( \lambda_{0} \right)$$

Then the Type I error $\alpha\left( \lambda_{0}\mathcal{,D} \right)$ is non-increasing on [$\lambda_{0L},\lambda_{0U}]$. If the above inequality is strict, then $\alpha\left( \lambda_{0}\mathcal{,D} \right)$ is strictly decreasing on [$\lambda_{0L},\lambda_{0U}]$.

**Remark:** this theorem provides a sufficient condition of monotonicity. The two terms on the left-hand side are the negative contributions to $\alpha'(\lambda_{0}\mathcal{,D)}$ induced by the monotone decrease of $\bar{c_{1}}(\lambda_{0})$ and $\bar{c_{2}}(\lambda_{0})$, and the term on the right-hand side is the positive contribution induced by the increase of $\rho_{H_{0}}\left( \lambda_{0} \right)$. In practice, this means that as the true hazard rate becomes less favorable, the design becomes less likely to reject the null, rather than becoming more likely to declare efficacy due to the increasing correlation between the interim ($Z_{1}$) and final test statistics ($Z_{2}$).

Proof: By chain rule,

$$\alpha'\left( \lambda_{0}\mathcal{,D} \right)=\left. \frac{\partial\Phi_{2}}{\partial a}\left( a,b;\rho\right) \right|_{a=\bar{c_{1}}(\lambda_{0}),b=\bar{c_{2}}(\lambda_{0}),\rho=\rho_{H_{0}}(\lambda_{0})}{\bar{c_{1}}}^{'}\left( \lambda_{0} \right)+\left. \frac{\partial\Phi_{2}}{\partial b}\left( a,b;\rho\right) \right|_{a=\bar{c_{1}}(\lambda_{0}),b=\bar{c_{2}}(\lambda_{0}),\rho=\rho_{H_{0}}(\lambda_{0})}{\bar{c_{2}}}^{'}\left( \lambda_{0} \right)+\left. \frac{\partial\Phi_{2}}{\partial\rho}\left( a,b;\rho\right) \right|_{a=\bar{c_{1}}(\lambda_{0}),b=\bar{c_{2}}(\lambda_{0}),\rho=\rho_{H_{0}}(\lambda_{0})}{\rho^{'}}_{H_{0}}\left( \lambda_{0} \right)=\phi\left( \bar{c_{1}}(\lambda_{0}) \right)\Phi\left( \frac{\bar{c_{2}}(\lambda_{0})-\bar{c_{1}}{(\lambda_{0})\rho}_{H_{0}}\left( \lambda_{0} \right)}{s\left( \lambda_{0} \right)} \right){\bar{c_{1}}}^{'}\left( \lambda_{0} \right)+\phi\left( \bar{c_{2}}(\lambda_{0}) \right)\Phi\left( \frac{\bar{c_{1}}(\lambda_{e})-\bar{c_{2}}{(\lambda_{e})\rho}_{H_{0}}\left( \lambda_{e} \right)}{s\left( \lambda_{e} \right)} \right){\bar{c_{2}}}^{'}\left( \lambda_{0} \right)+\phi_{2}\left( \bar{c_{1}}\left( \lambda_{0} \right),\bar{c_{2}}\left( \lambda_{0} \right);\rho_{H_{0}}\left( \lambda_{0} \right) \right){\rho^{'}}_{H_{0}}\left( \lambda_{0} \right)$$

Since ${\bar{c_{k}}}^{'}\left( \lambda_{0} \right)<0$ for $k=1,2$, from the assumption, we have $\alpha^{'}\left( \lambda_{0}\mathcal{,D} \right)\leq0$. And whenever the inequality in assumption is strict, we have $\alpha^{'}\left( \lambda_{0}\mathcal{,D} \right)<0$.

**Corollary 1.** Under the assumptions of theorem 1, the worst-case type I error over the hazard rate benchmark range $H_{0}$ is attained at the left endpoint:

$$\sup_{\lambda_{0}\in[\lambda_{0L},\lambda_{0U}]} \alpha(\lambda_{0}\mathcal{,D)}=\alpha(\lambda_{0L}\mathcal{,D)}$$

**Corollary 2.** Under the assumptions of theorem 1, let $S=e^{-\lambda x_{0}}$ denote the survival probability at time $x_{0}$, then the worst-case type I error over the survival probability benchmark range $H_{0}$ is attained at the right endpoint:

$$\sup_{s\in[S_{0L},S_{0U}]} \alpha(s,\mathcal{D)}=\alpha(S_{0U}\mathcal{,D)}$$

Proof: the result follows as the map $\lambda\mapsto S=e^{-\lambda x_{0}}$ is strictly decreasing.

**Remark:** The well-defineness arguments for the BUDS objectives extend directly to the TTE setting. For any fixed r-KJ design $d=\left( n_{1},c_{1},DA_{1};n_{2},c_{2},DA_{2} \right)$, the expected sample size can be written as $EN_{d}\left( S \right)=n_{1}+\left( 1-PET_{d}\left( S \right) \right)\left( n_{2}-n_{1} \right)$, and $PET_{d}\left( S \right)$ follows the same definition as in binary endpoint case. Thus, regret is finite over the benchmark range $\left[ S_{0L},S_{0U} \right]$. Because the candidate design space $\mathcal{D}$ is finite for any prespecified $n_{max}$ , the maximum regret and the BUDS-Worst-Regret minimizer $d_{WR}$ are well-defined whenever the feasible set is non-empty.
