## Supplementary Material 3 for "BUDS: Benchmark Uncertainty Design Selection for Two-Stage Single-Arm Phase II Trials"

**Supplementary Tables and Figures**

**Table S1:** Design Boundaries and Exact Operating Characteristics Under Benchmark Uncertainty for Binary Endpoints ($\alpha$ = 0.05, $\beta$= 0.20, $n_{\max}$ = 150)

| **Benchmark Range**  $\boldsymbol{[}\boldsymbol{p}_{\boldsymbol{0}\boldsymbol{L}}\boldsymbol{,}\boldsymbol{p}_{\boldsymbol{0}\boldsymbol{U}}\boldsymbol{]}$ | **Design** | **Boundaries**  $\boldsymbol{(}\boldsymbol{n}_{\boldsymbol{1}}\boldsymbol{,}\boldsymbol{r}_{\boldsymbol{1}}\boldsymbol{;n, r)}$ | **Avg. EN over** $\boldsymbol{[}\boldsymbol{p}_{\boldsymbol{0}\boldsymbol{L}}\boldsymbol{,}\boldsymbol{p}_{\boldsymbol{0}\boldsymbol{U}}\boldsymbol{]}$ | **Power at** $\boldsymbol{p}_{\boldsymbol{1}}$ | **Type I Error at** $\boldsymbol{[}\boldsymbol{p}_{\boldsymbol{0}\boldsymbol{L}}\boldsymbol{,}\boldsymbol{p}_{\boldsymbol{0}\boldsymbol{U}}\boldsymbol{]}$ |
| --- | --- | --- | --- | --- | --- |
| **Scenario 1:**$\boldsymbol{p}_{\boldsymbol{1}}$ **= 0.25** | | | | | |
| [0.10, 0.10] | Optimal | (18, 2; 43, 7) | 24.7 | 0.800 | [0.048, 0.048] |
|  | Minimax | (22, 2; 40, 7) | 28.8 | 0.803 | [0.040, 0.040] |
|  | BUDS-Worst-Regret | (18, 2; 43, 7) | 24.7 | 0.800 | [0.048, 0.048] |
| [0.08, 0.12] | Optimal – $p_{0}$ | (18, 2; 43, 7) | 24.7 | 0.800 | [0.015, 0.110] |
|  | Optimal – $p_{0U}$ | (19, 2; 61, 11) | 31.4 | 0.803 | [0.002, 0.048] |
|  | Minimax – $p_{0}$ | (22, 2; 40, 7) | 28.8 | 0.803 | [0.012, 0.095] |
|  | Minimax – $p_{0U}$ | (30, 3; 53, 10) | 38.1 | 0.801 | [0.003, 0.046] |
|  | BUDS-Worst-Regret | (18, 2; 68, 12) | 31.4 | 0.805 | [0.002, 0.046] |
| [0.05, 0.15] | Optimal – $p_{0}$ | (18, 2; 43, 7) | 24.8 | 0.800 | [0.001, 0.258] |
|  | Optimal – $p_{0U}$ | (41, 7; 129, 25) | 47.9 | 0.800 | [<0.001, 0.048] |
|  | Minimax – $p_{0}$ | (22, 2; 40, 7) | 28.8 | 0.803 | [0.001, 0.234] |
|  | Minimax – $p_{0U}$ | (55, 8; 97, 20) | 61.0 | 0.801 | [<0.001, 0.049] |
|  | BUDS-Worst-Regret | (28, 4; 122, 24) | 43.7 | 0.801 | [<0.001, 0.049] |
| **Scenario 2:** $\boldsymbol{p}_{\boldsymbol{1}}$ **= 0.40** | | | | | |
| [0.20, 0.20] | Optimal | (13, 3; 43, 12) | 20.6 | 0.800 | [0.050, 0.050] |
|  | Minimax | (18, 4; 33, 10) | 22.2 | 0.801 | [0.046, 0.046] |
|  | BUDS-Worst-Regret | (13, 3; 43, 12) | 20.6 | 0.800 | [0.050, 0.050] |
| [0.18, 0.22] | Optimal – $p_{0}$ | (13, 3; 43, 12) | 20.6 | 0.800 | [0.024, 0.091] |
|  | Optimal – $p_{0U}$ | (19, 5; 54, 16) | 24.8 | 0.804 | [0.008, 0.047] |
|  | Minimax – $p_{0}$ | (18, 4; 33, 10) | 22.3 | 0.801 | [0.023, 0.082] |
|  | Minimax – $p_{0U}$ | (22, 5; 41, 13) | 27.1 | 0.803 | [0.009, 0.047] |
|  | BUDS-Worst-Regret | (13, 3; 58, 17) | 24.4 | 0.800 | [0.007, 0.048] |
| [0.15, 0.25] | Optimal – $p_{0}$ | (13, 3; 43, 12) | 20.7 | 0.800 | [0.005, 0.186] |
|  | Optimal – $p_{0U}$ | (20, 5; 71, 23) | 30.5 | 0.802 | [<0.001, 0.049] |
|  | Minimax – $p_{0}$ | (18, 4; 33, 10) | 22.3 | 0.801 | [0.006, 0.166] |
|  | Minimax – $p_{0U}$ | (51, 16; 60, 20) | 51.3 | 0.803 | [<0.001, 0.050] |
|  | BUDS-Worst-Regret | (20, 5; 71, 23) | 30.5 | 0.802 | [<0.001, 0.049] |
| **Scenario 3:** $\boldsymbol{p}_{\boldsymbol{1}}$ **= 0.50** | | | | | |
| [0.30, 0.30] | Optimal | (15, 5; 46, 18) | 23.6 | 0.803 | [0.050, 0.050] |
|  | Minimax | (19, 6; 39, 16) | 25.7 | 0.804 | [0.046, 0.046] |
|  | BUDS-Worst-Regret | (15, 5; 46, 18) | 23.6 | 0.803 | [0.050, 0.050] |
| [0.28, 0.32] | Optimal – $p_{0}$ | (15, 5; 46, 18) | 23.6 | 0.803 | [0.026, 0.087] |
|  | Optimal – $p_{0U}$ | (20, 7; 56, 23) | 28.2 | 0.805 | [0.009, 0.045] |
|  | Minimax – $p_{0}$ | (19, 6; 39, 16) | 25.7 | 0.804 | [0.024, 0.079] |
|  | Minimax – $p_{0U}$ | (21, 6; 47, 20) | 32.7 | 0.800 | [0.010, 0.045] |
|  | BUDS-Worst-Regret | (17, 6; 65, 26) | 27.8 | 0.801 | [0.009, 0.046] |
| [0.25, 0.35] | Optimal – $p_{0}$ | (15, 5; 46, 18) | 23.8 | 0.803 | [0.008, 0.172] |
|  | Optimal – $p_{0U}$ | (27, 10; 77, 33) | 35.4 | 0.801 | [<0.001, 0.049] |
|  | Minimax – $p_{0}$ | (19, 6; 39, 16) | 25.8 | 0.804 | [0.008, 0.156] |
|  | Minimax – $p_{0U}$ | (55, 22; 66, 29) | 55.6 | 0.801 | [<0.001, 0.050] |
|  | BUDS-Worst-Regret | (22, 8; 85, 36) | 34.4 | 0.802 | [<0.001, 0.049] |

Design boundaries and exact operating characteristics for Simon Optimal, Simon Minimax, and BUDS-Worst-Regret-selected designs under varying levels of benchmark uncertainty. Results are shown for three scenarios defined by $(p_{0}, p_{1})$. For each benchmark range $[p_{0L}, p_{0U}]$, the table reports the design boundaries $(n_{1},r_{1};n, r)$, the average expected sample size over the range, statistical power at $p_{1}$, and the range of Type I error probabilities across $[p_{0L}, p_{0U}]$. For comparison, the Optimal and Minimax designs are shown for planning benchmarks of $p_{0}$ and $p_{0U}$.

**Table S2:** Design Boundaries and Exact Operating Characteristics Under Benchmark Uncertainty for Time-to-Event Endpoints ($\alpha$ = 0.10, $\beta$= 0.10, $n_{\max}$ = 150)

| **Benchmark**  **Range** $\boldsymbol{[}\boldsymbol{S}_{\boldsymbol{0}\boldsymbol{L}}\boldsymbol{(}\boldsymbol{x}_{\boldsymbol{0}}\boldsymbol{)}\boldsymbol{,}\boldsymbol{S}_{\boldsymbol{0}\boldsymbol{U}}\boldsymbol{(}\boldsymbol{x}_{\boldsymbol{0}}\boldsymbol{)]}$ | **Design** | **Calibration Benchmark** $\boldsymbol{S}_{\boldsymbol{0}}^{\mathbf{cal}}\left( \boldsymbol{x}_{\boldsymbol{0}} \right)$ | **Boundaries**  $\left( \begin{aligned} \boldsymbol{n}_{\boldsymbol{1}}\boldsymbol{,}\boldsymbol{c}_{\boldsymbol{1}}\boldsymbol{,D}\boldsymbol{A}_{\boldsymbol{1}}\boldsymbol{;} \\ \boldsymbol{n}_{\boldsymbol{2}}\boldsymbol{,}\boldsymbol{c}_{\boldsymbol{2}}\boldsymbol{,D}\boldsymbol{A}_{\boldsymbol{2}} \end{aligned} \right)$ | **Avg. EN over** $\boldsymbol{[}\boldsymbol{S}_{\boldsymbol{0}\boldsymbol{L}}\boldsymbol{(}\boldsymbol{x}_{\boldsymbol{0}}\boldsymbol{)}\boldsymbol{,}\boldsymbol{S}_{\boldsymbol{0}\boldsymbol{U}}\boldsymbol{(}\boldsymbol{x}_{\boldsymbol{0}}\boldsymbol{)]}$ | **Power at** $\boldsymbol{S}_{\boldsymbol{1}}\boldsymbol{(}\boldsymbol{x}_{\boldsymbol{0}}\boldsymbol{)}$ | **Type I error at** $\boldsymbol{[}\boldsymbol{S}_{\boldsymbol{0}\boldsymbol{L}}\boldsymbol{(}\boldsymbol{x}_{\boldsymbol{0}}\boldsymbol{)}\boldsymbol{,}\boldsymbol{S}_{\boldsymbol{0}\boldsymbol{U}}\boldsymbol{(}\boldsymbol{x}_{\boldsymbol{0}}\boldsymbol{)]}$ |
| --- | --- | --- | --- | --- | --- | --- |
| **Scenario 1:** $\boldsymbol{S}_{\boldsymbol{1}}\boldsymbol{(}\boldsymbol{x}_{\boldsymbol{0}}\boldsymbol{)}\mathbf{= 0.34}$ | | | | | | |
| [0.15, 0.15] | r-KJ | 0.15 | (20, 0.73, 0.64;  26, -1.27, 1.87) | 24.6 | 0.901 | [0.100, 0.100] |
|  | BUDS-Worst-Regret | 0.15 | (20, 0.73, 0.64;  26, -1.27, 1.87) | 24.6 | 0.901 | [0.100, 0.100] |
| [0.13, 0.17] | r-KJ – $S_{0}(x_{0})$ | 0.15 | (20, 0.73, 0.64;  26, -1.27, 1.87) | 24.6 | 0.901 | [0.058, 0.159] |
|  | r-KJ – $S_{0U}(x_{0})$ | 0.17 | (26, 0.45, 0.86;  35, -1.27, 2.17) | 31.3 | 0.900 | [0.028, 0.100] |
|  | BUDS-Worst-Regret | 0.17 | (24, 0.50, 0.80;  36, -1.26, 2.20) | 31.4 | 0.902 | [0.027, 0.100] |
| [0.10, 0.20] | r-KJ – $S_{0}(x_{0})$ | 0.15 | (20, 0.73, 0.64;  26, -1.27, 1.87) | 24.5 | 0.901 | [0.022, 0.282] |
|  | r-KJ – $S_{0U}(x_{0})$ | 0.20 | (40, 0.25, 1.30;  57, -1.27, 2.90) | 46.0 | 0.901 | [0.001, 0.100] |
|  | BUDS-Worst-Regret | 0.20 | (38, 0.36, 1.27;  57, -1.27, 2.90) | 45.6 | 0.901 | [0.001, 0.100] |
| **Scenario 2:** $\boldsymbol{S}_{\boldsymbol{1}}\boldsymbol{(}\boldsymbol{x}_{\boldsymbol{0}}\boldsymbol{)}\mathbf{= 0.59}$ | | | | | | |
| [0.35, 0.35] | r-KJ | 0.35 | (20, 0.80, 0.66;  27, -1.27, 1.90) | 25.5 | 0.900 | [0.100, 0.100] |
|  | BUDS-Worst-Regret | 0.35 | (20, 0.80, 0.66;  27, -1.27, 1.90) | 25.5 | 0.900 | [0.100, 0.100] |
| [0.33, 0.37] | r-KJ – $S_{0}(x_{0})$ | 0.35 | (20, 0.80, 0.66;  27, -1.27, 1.90) | 25.5 | 0.900 | [0.070, 0.139] |
|  | r-KJ – $S_{0U}(x_{0})$ | 0.37 | (24, 0.53, 0.79;  33, -1.26, 2.10) | 29.9 | 0.900 | [0.045, 0.100] |
|  | BUDS-Worst-Regret | 0.37 | (22, 0.79, 0.73;  33, -1.27, 2.10) | 30.2 | 0.902 | [0.045, 0.100] |
| [0.30, 0.40] | r-KJ – $S_{0}(x_{0})$ | 0.35 | (20, 0.80, 0.66;  27, -1.27, 1.90) | 25.5 | 0.900 | [0.040, 0.216] |
|  | r-KJ – $S_{0U}(x_{0})$ | 0.40 | (33, 0.31, 1.07;  45, -1.26, 2.50) | 38.5 | 0.901 | [0.007, 0.100] |
|  | BUDS-Worst-Regret | 0.40 | (30, 0.50, 1.00;  45, -1.27, 2.50) | 38.2 | 0.901 | [0.007, 0.100] |
| **Scenario 3:**$\boldsymbol{S}_{\boldsymbol{1}}\boldsymbol{(}\boldsymbol{x}_{\boldsymbol{0}}\boldsymbol{)}\mathbf{= 0.67}$ | | | | | | |
| [0.50, 0.50] | r-KJ | 0.50 | (40, 0.26, 1.30;  56, -1.26, 2.87) | 49.6 | 0.901 | [0.100, 0.100] |
|  | BUDS-Worst-Regret | 0.50 | (40, 0.26, 1.30;  56, -1.26, 2.87) | 49.6 | 0.901 | [0.100, 0.100] |
| [0.48, 0.52] | r-KJ – $S_{0}(x_{0})$ | 0.50 | (40, 0.26, 1.30;  56, -1.26, 2.87) | 49.6 | 0.901 | [0.061, 0.156] |
|  | r-KJ – $S_{0U}(x_{0})$ | 0.52 | (50, 0.19, 1.64;  71, -1.26, 3.37) | 60.4 | 0.900 | [0.031, 0.100] |
|  | BUDS-Worst-Regret | 0.52 | (48, 0.36, 1.58;  71, -1.27, 3.37) | 60.9 | 0.902 | [0.031, 0.100] |
| [0.45, 0.55] | r-KJ – $S_{0}(x_{0})$ | 0.50 | (40, 0.26, 1.30;  56, -1.26, 2.87) | 49.6 | 0.901 | [0.027, 0.280] |
|  | r-KJ – $S_{0U}(x_{0})$ | 0.55 | (74, 0.11, 2.45;  109, -1.27, 4.63) | 85.0 | 0.900 | [0.001, 0.100] |
|  | BUDS-Worst-Regret | 0.55 | (55, 0.50, 1.80;  116, -1.26, 4.87) | 86.5 | 0.901 | [0.001, 0.100] |

Design boundaries and exact operating characteristics for restricted-KJ and BUDS-Worst-Regret-selected designs under varying levels of benchmark uncertainty. Results are shown for three scenarios defined by $(S_{0}\left( x_{0} \right), S_{1}\left( x_{0} \right))$. For each benchmark range $[S_{0L}(x_{0}), S_{0U}(x_{0})]$, the table reports the design boundaries $(c_{1},n_{1}, DA_{1};c_{2}, n_{2},DA_{2})$, the average expected sample size over the range, statistical power at $S_{1}(x_{0})$, and the range of Type I error probabilities across $[S_{0L}(x_{0}), S_{0U}(x_{0})]$. The column The column *Calibration Benchmark* $S_{0}^{\mathrm{cal}}\left( x_{0} \right)$ denotes the evaluation benchmark survival probability at which the design selected by the objective is constructed. For comparison, the r-KJ design is shown for planning benchmarks of $S_{0}(x_{0})$ and $S_{0U}(x_{0}$), with the target survival probability $S_{1}\left( x_{0} \right))$ held fixed in both cases (resulting in different derived hazard ratios). Design inputs: $x_{0}$ = 1, accrual rate = 30.

**Figure S1:** Type I Error Rate as a Function of the True Benchmark by Design

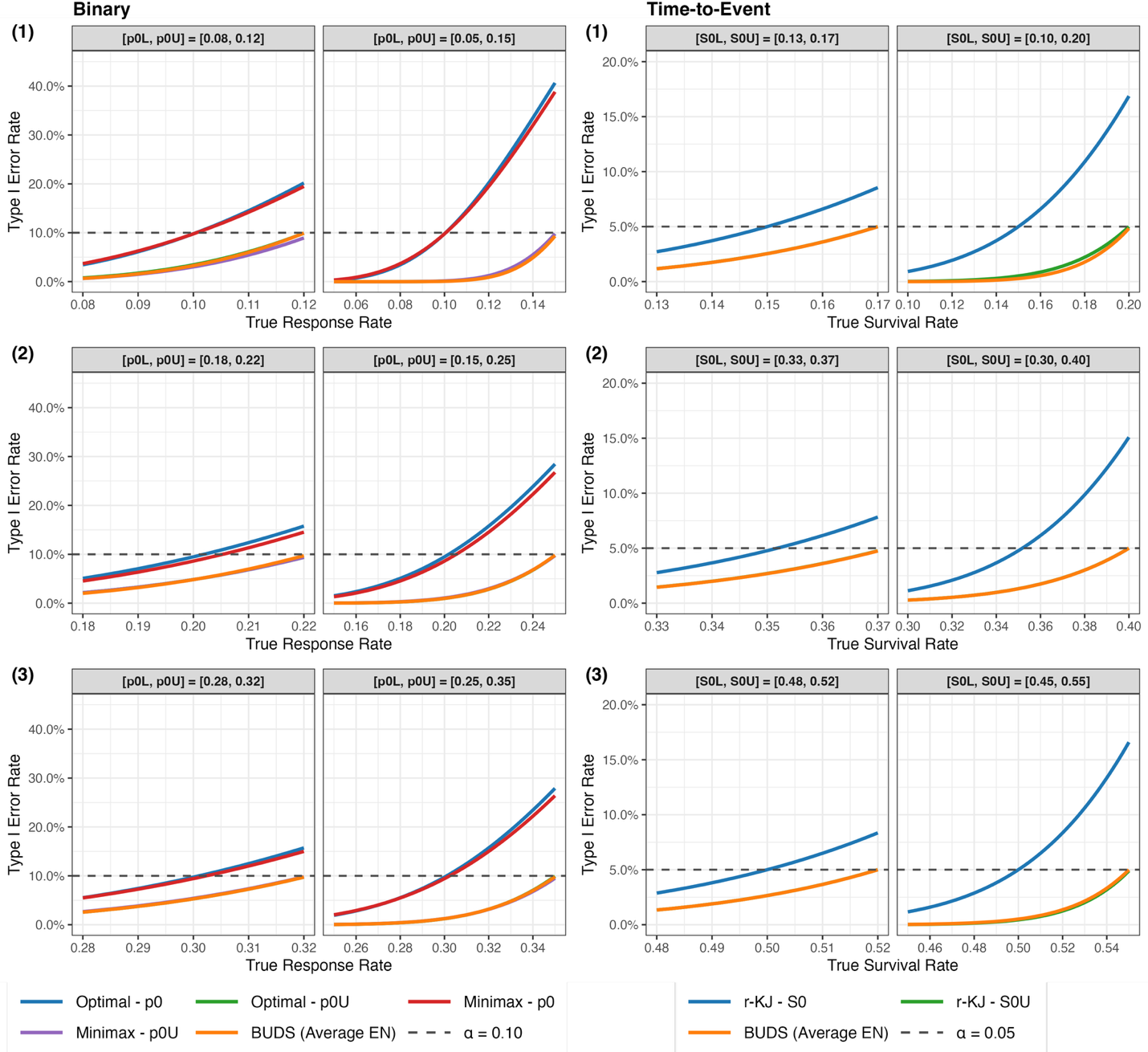

**Legend:** Type I error curves for designs selected by single-benchmark and BUDS objectives across the benchmark range. Panel (A) shows binary endpoint scenarios with $(p_{0},p_{1})$ = (0.10, 0.25), (0.20, 0.40), and (0.30, 0.50), planned with $\alpha$ = 0.10, $\beta$ = 0.10, and $n_{max}$ = 150. Panel (B) shows time-to-event scenarios defined by $(S_{0}\left( x_{0} \right), S_{1}\left( x_{0} \right)$) = (0.15, 0.34), (0.35, 0.59), and (0.50, 0.67) at $x_{0}$ = 1, planned with $\alpha$ = 0.05, $\beta$ = 0.20, a uniform accrual rate of 30, and $n_{max}$ = 150. The dashed horizontal lines denote the nominal Type I error level. Within each scenario, results are shown for two benchmark ranges $[\theta_{0L}, \theta_{0U}]$, representing moderate and substantial uncertainty. For comparison, the single-benchmark designs are shown for planning benchmarks of both $\theta_{0}$ and $\theta_{0U}$. The BUDS-Average-EN-selected designs maintain Type I error rate at or below $\alpha$ throughout the benchmark range, whereas classic designs calibrated at $\theta_{0}$ exhibit increasing Type I error rate as the evaluation benchmark approaches the upper boundary of the range. Because Type I error is monotone in the benchmark, designs calibrated at $\theta_{0U}$ also control Type I error across the benchmark range; consequently, some BUDS and $\theta_{0U}$-calibrated designs coincide.

**Table S3:** Sensitivity of BUDS-Worst-Regret Designs to the Specified Alternative Response Rate Under Benchmark Uncertainty for Binary Endpoints ($n_{\max}$ = 150)

| $\boldsymbol{p}_{\boldsymbol{0}}$ | $\boldsymbol{[}\boldsymbol{p}_{\boldsymbol{0}\boldsymbol{L}}\boldsymbol{,}\boldsymbol{p}_{\boldsymbol{0}\boldsymbol{U}}\boldsymbol{]}$ | $\boldsymbol{p}_{\boldsymbol{1}}$ | **Boundaries**  $\boldsymbol{(}\boldsymbol{n}_{\boldsymbol{1}}\boldsymbol{,}\boldsymbol{r}_{\boldsymbol{1}}\boldsymbol{;n, r)}$ | **Avg. EN over** $\boldsymbol{[}\boldsymbol{p}_{\boldsymbol{0}\boldsymbol{L}}\boldsymbol{,}\boldsymbol{p}_{\boldsymbol{0}\boldsymbol{U}}\boldsymbol{]}$ | **Power at** $\boldsymbol{p}_{\boldsymbol{1}}$ | **Type I error at** $\boldsymbol{[}\boldsymbol{p}_{\boldsymbol{0}\boldsymbol{L}}\boldsymbol{,}\boldsymbol{p}_{\boldsymbol{0}\boldsymbol{U}}\boldsymbol{]}$ |
| --- | --- | --- | --- | --- | --- | --- |
| $\boldsymbol{\alpha}$**= 0.10,** $\boldsymbol{\beta}$ **= 0.10** | | | | | | |
| 0.05 | [0.05, 0.05] | 0.15 | (28, 1; 66, 5) | 43.64 | 0.902 | [0.095, 0.095] |
|  |  | 0.20 | (12, 0; 37, 3) | 23.49 | 0.902 | [0.094, 0.094] |
|  |  | 0.25 | (9, 0; 24, 2) | 14.55 | 0.903 | [0.093, 0.093] |
|  | [0.03, 0.07] | 0.15 | (39, 2; 115, 11) | 62.66 | 0.902 | [<0.001, 0.095] |
|  |  | 0.20 | (22, 1; 47, 5) | 29.54 | 0.903 | [0.002, 0.098] |
|  |  | 0.25 | (12, 0; 26, 3) | 18.36 | 0.905 | [0.007, 0.099] |
|  | [0.00, 0.10] | 0.15 | No feasible design | - | - | - |
|  |  | 0.20 | (21, 1; 105, 14) | 45.37 | 0.902 | [<0.001, 0.089] |
|  |  | 0.25 | (10, 0; 49, 7) | 24.65 | 0.903 | [<0.001, 0.099] |
| 0.10 | [0.10, 0.10] | 0.20 | (41, 4; 99, 13) | 63.71 | 0.903 | [0.097, 0.097] |
|  |  | 0.25 | (21, 2; 50, 7) | 31.20 | 0.901 | [0.098, 0.098] |
|  |  | 0.30 | (12, 1; 35, 5) | 19.84 | 0.901 | [0.098, 0.098] |
|  | [0.08, 0.12] | 0.20 | (65, 7; 146, 22) | 91.54 | 0.903 | [0.001, 0.098] |
|  |  | 0.25 | (27, 3; 69, 11) | 38.93 | 0.903 | [0.007, 0.099] |
|  |  | 0.30 | (17, 2; 42, 7) | 22.99 | 0.903 | [0.013, 0.096] |
|  | [0.05, 0.15] | 0.20 | No feasible design | - | - | - |
|  |  | 0.25 | (34, 4; 118, 22) | 56.72 | 0.901 | [<0.001, 0.099] |
|  |  | 0.30 | (20, 2; 54, 11) | 31.11 | 0.902 | [<0.001, 0.095] |
| $\boldsymbol{\alpha}$**= 0.05,** $\boldsymbol{\beta}$ **= 0.20** | | | | | | |
| 0.05 | [0.05, 0.05] | 0.15 | (23, 1; 56, 5) | 33.58 | 0.800 | [0.050, 0.050] |
|  |  | 0.20 | (10, 0; 29, 3) | 17.62 | 0.801 | [0.047, 0.047] |
|  |  | 0.25 | (9, 0; 17, 2) | 11.96 | 0.812 | [0.047, 0.047] |
|  | [0.03, 0.07] | 0.15 | (31, 2; 115, 12) | 48.35 | 0.802 | [<0.001, 0.046] |
|  |  | 0.20 | (16, 1; 50, 6) | 22.51 | 0.803 | [0.001, 0.045] |
|  |  | 0.25 | (7, 0; 30, 4) | 13.89 | 0.806 | [0.001, 0.044] |
|  | [0.00, 0.10] | 0.15 | No feasible design | - | - | - |
|  |  | 0.20 | (16, 1; 105, 15) | 34.28 | 0.803 | [<0.001, 0.045] |
|  |  | 0.25 | (9, 0; 42, 7) | 20.49 | 0.813 | [<0.001, 0.049] |
| 0.10 | [0.10, 0.10] | 0.20 | (30, 3; 89, 13) | 50.80 | 0.802 | [0.048, 0.048] |
|  |  | 0.25 | (18, 2; 43, 7) | 24.66 | 0.800 | [0.048, 0.048] |
|  |  | 0.30 | (10, 1; 29, 5) | 15.01 | 0.805 | [0.047, 0.047] |
|  | [0.08, 0.12] | 0.20 | (49, 7; 144, 23) | 70.01 | 0.801 | [<0.001, 0.050] |
|  |  | 0.25 | (18, 2; 68, 12) | 31.37 | 0.805 | [0.002, 0.046] |
|  |  | 0.30 | (11, 1; 35, 7) | 18.26 | 0.802 | [0.005, 0.043] |
|  | [0.05, 0.15] | 0.20 | No feasible design | - | - | - |
|  |  | 0.25 | (28, 4; 122, 24) | 43.71 | 0.801 | [<0.001, 0.049] |
|  |  | 0.30 | (15, 2; 59, 13) | 23.58 | 0.801 | [<0.001, 0.041] |

Sensitivity of BUDS-Worst-Regret designs to the specified alternative response rate under benchmark uncertainty for representative binary scenarios. For each benchmark range $[p_{0L}, p_{0U}]$, the table reports the design boundaries $(n_{1},r_{1};n, r)$, the average expected sample size over the range, statistical power at $p_{1}$, and the range of Type I error probabilities across $[p_{0L}, p_{0U}]$. “No feasible design” indicates that no design satisfying the prespecified Type I error and power constraints was identified within the maximum allowable sample size ($n_{\max}$).
