## Supplementary Material 4 for "BUDS: Benchmark Uncertainty Design Selection for Two-Stage Single-Arm Phase II Trials"

**Shiny Application Workflow**

The accompanying Shiny application provides an interactive interface for constructing two-stage trial designs under BUDS, visualizing their operating characteristics, and generating downloadable reports.

**Binary Endpoints**

The workflow proceeds as follows:

1. **Specify design parameters**
   Users begin by entering the required inputs in the interface, including the benchmark range $[p_{0L},p_{0U}]$, target response rate $p_{1}$, error constraints (Type I error $\alpha$ and power), and the maximum sample size $n_{\max}$. The desired BUDS objective(s) (Worst-Regret or Average-EN) can also be selected.
2. **Run the design search**
   After specifying inputs, users click *Run*, which initiates the design search procedure. The default output is displayed in the *Search Results* view, presenting a summary table of candidate designs and their key operating characteristics.
3. **Navigate results**
   Users can explore outputs using the results navigation panel:
   - **Search Results**: displays candidate designs and summary metrics.

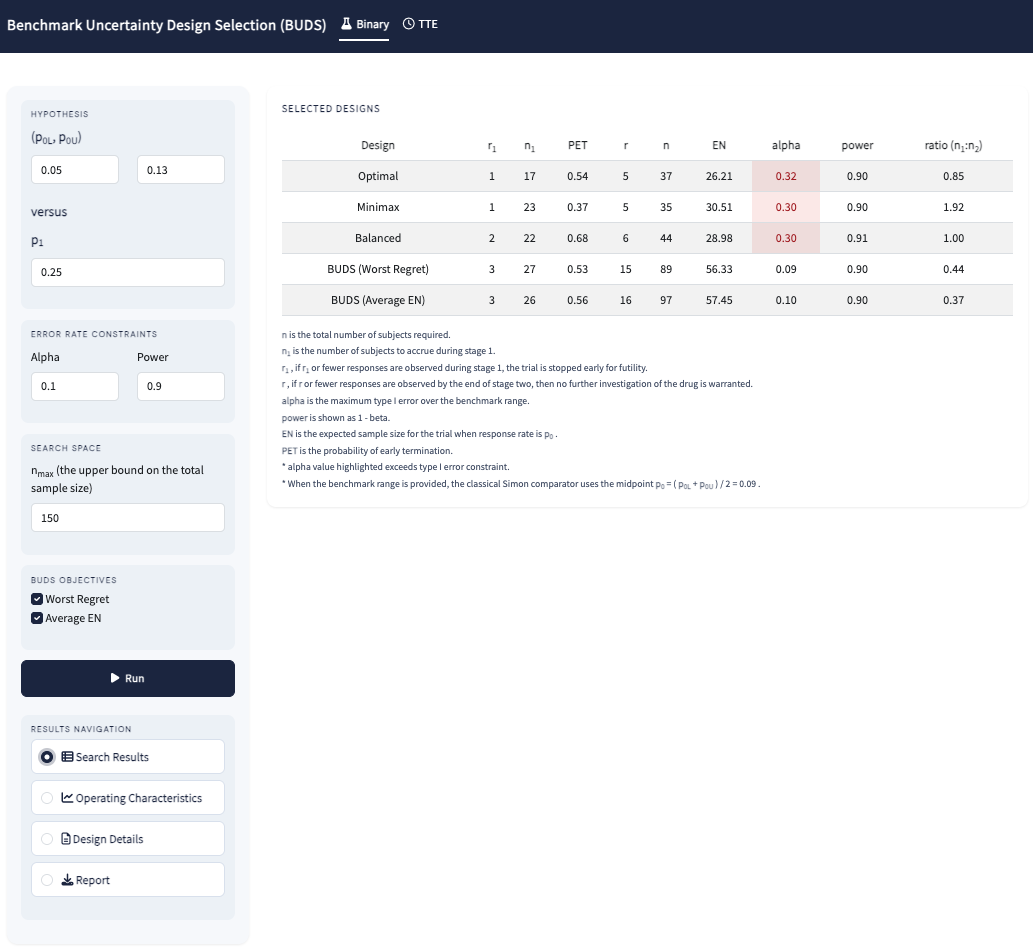

- **Operating Characteristics**: provides a detailed table of operating characteristics across the benchmark range, along with a Type I error rate plot.

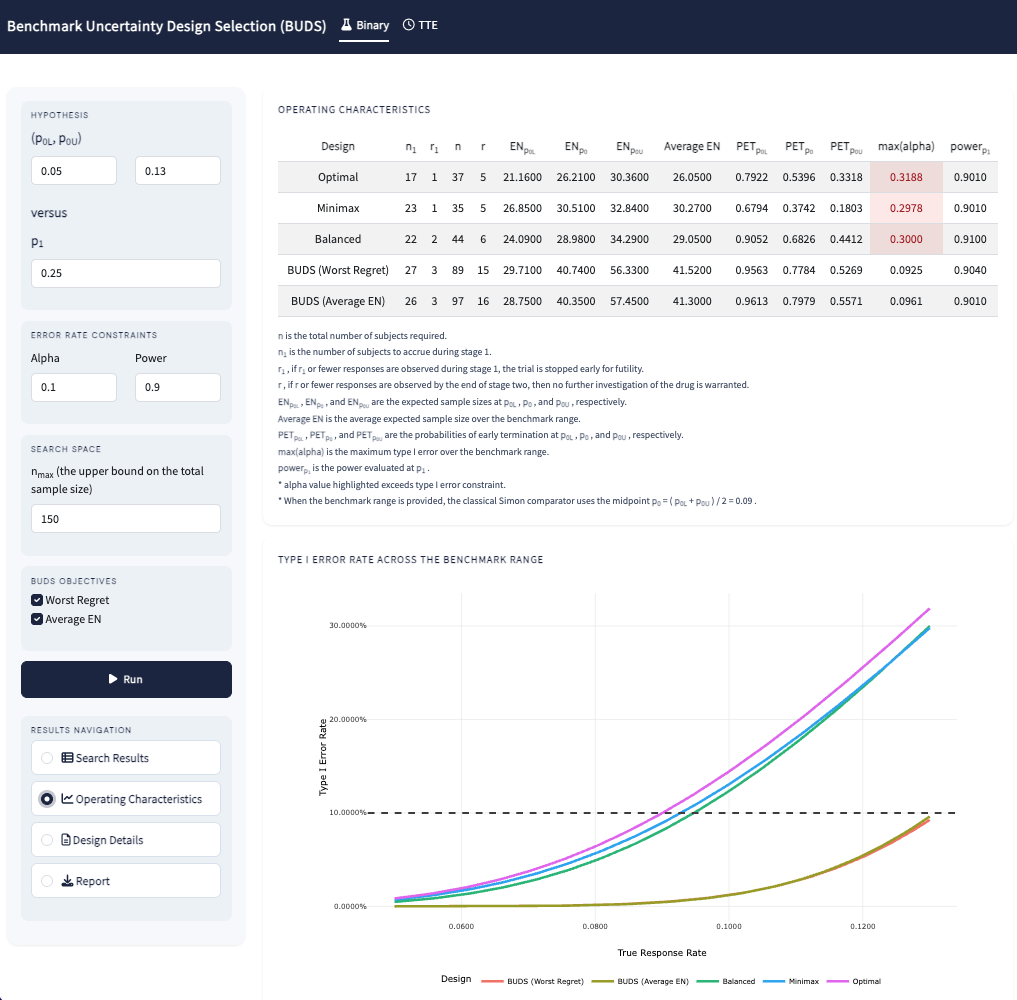

- **Design Details**: displays detailed information for a selected design, including stage-specific decision rules, sample sizes, and summaries of operating characteristics.

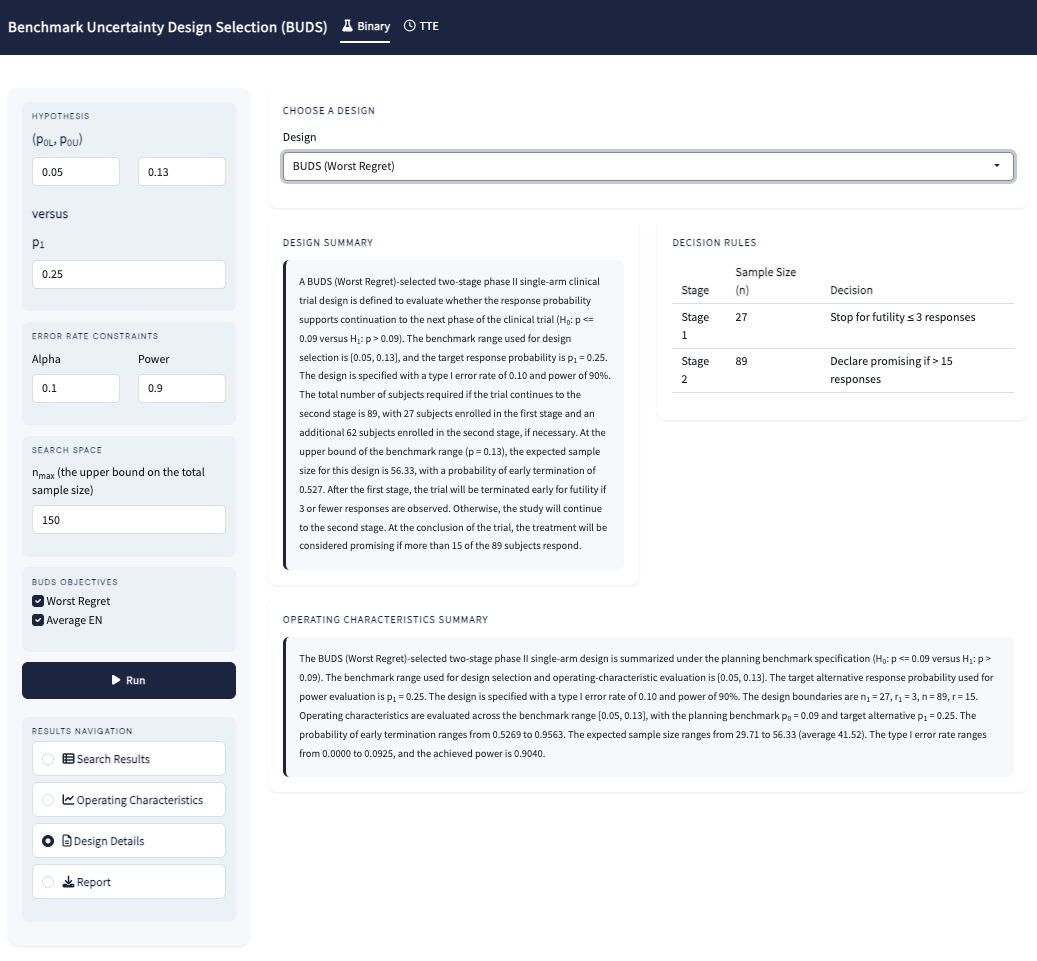

4. **Export results**
   Users can generate a downloadable HTML report containing all outputs, including summary tables, operating characteristics, plots, and detailed design summaries for selected designs.

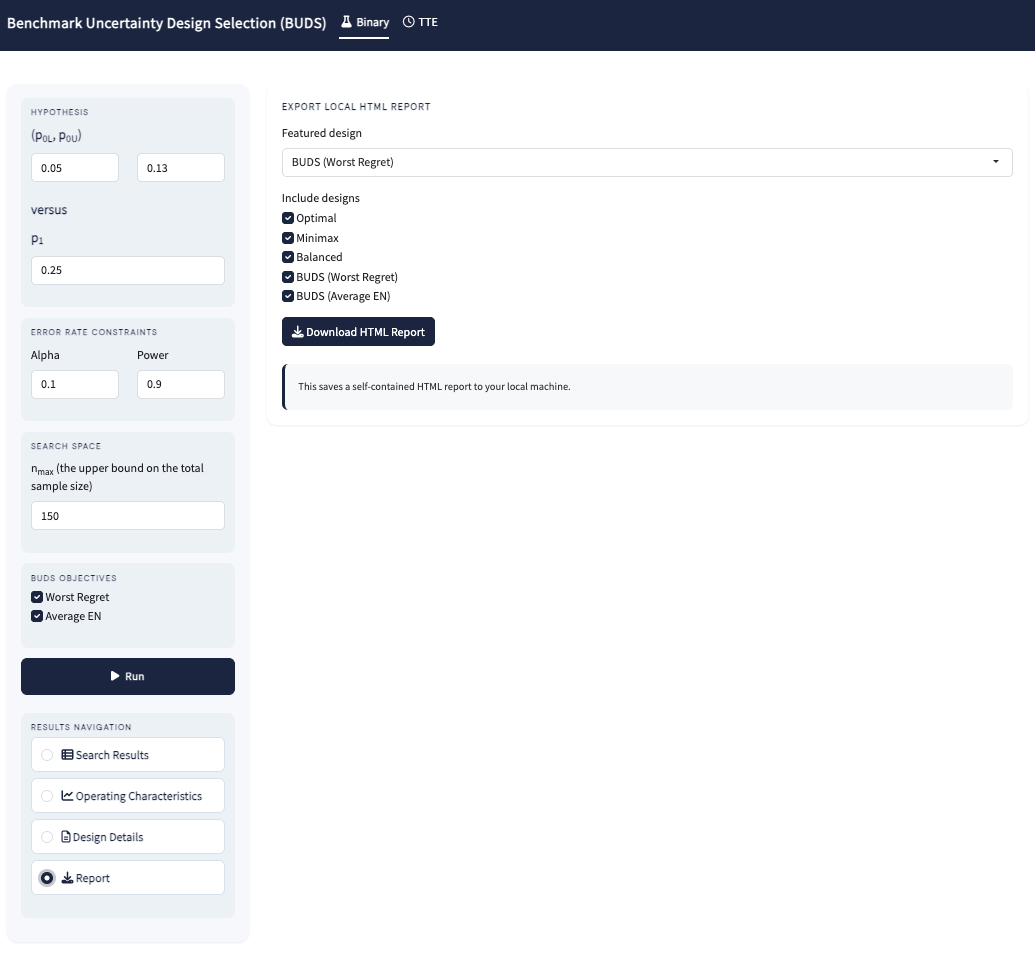

**Time-to-event Endpoints**

The workflow is identical to that described for binary endpoints, with the only difference being the specification of input parameters.

1. **Specify design parameters**
   Users begin by entering the required inputs in the interface, including the benchmark range using either survival probability $[S_{0L}\left( x_{0} \right),S_{0U}\left( x_{0} \right)]$ or hazard rate $[\lambda_{0L},\lambda_{0U}]$. The alternative can be specified via the target survival probability $S_{1}(x_{0})$, hazard ratio (HR), or alternative hazard $\lambda_{1}$, depending on the selected input type. Additional inputs include the restricted follow-up time $x_{0}$, accrual rate, error constraints (Type I error $\alpha$ and power), and the maximum sample size $n_{\max}$. The desired BUDS objective(s) (Worst-Regret or Average EN) can also be selected.

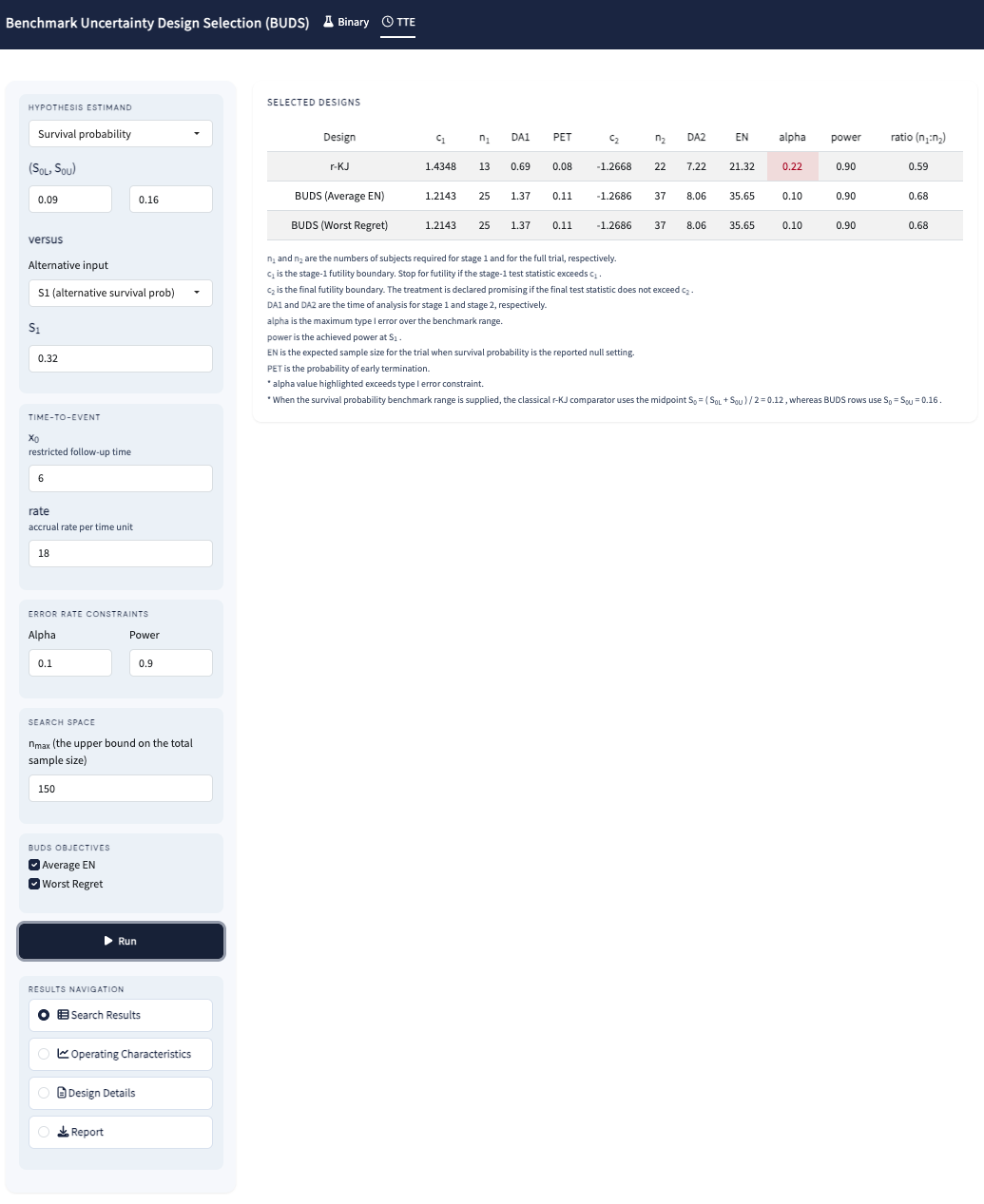
